# Bayesian inference of genetic pleiotropy identifies drug targets and repurposable medicines for human complex diseases

**DOI:** 10.1101/2025.03.17.25324106

**Authors:** Noah Lorincz-Comi, Feixiong Cheng

## Abstract

Complex diseases share heritable components which can be leveraged to identify drug targets with low side effect or high repurposing potential, but current methods cannot efficiently make these inferences at scale using public data. We introduce a Bayesian model to estimate the polygenic structure of a trait using GWAS summary data (BPACT). Across 32 complex traits, we estimated that 69.5 to 97.5% of disease-associated druggable genes are shared between multiple traits. We observed that targeting *KIT* for ALS prevention may increase triglyceride levels, but that targeting *TBK1* and *SCN11B* may be safer because of they were not pleiotropic. We additionally found 21 candidate repurposable drug targets for Alzheimer’s disease (AD) (e.g., *PLEKHA1, PPIB*) and 5 for ALS (e.g., *GAK, DGKQ*) based on the directionality of their pleiotropy. Our results demonstrate that modeling shared genetic architecture across traits can uncover safer therapeutic targets and highlight opportunities for drug repurposing in complex diseases.

**Motivation:** We intend to identify genes which are associated with multiple complex traits such that their side effect or therapeutic repurposing potential is large. We present a Bayesian method leveraging gene-based association test statistics that can be used to jointly characterize shared polygenicity between complex trait pairs and make these inferences.

## Introduction

Understanding the extent to which the heritability of a complex trait is conferred by few or many genes, i.e., its ‘polygenicity’, is a first step in understanding its genetic etiology [1]. Similarly, quantifying the degree to which two distinct traits have heritability which is contributed by shared and non-shared genes helps researchers better contextualize phenotypic similarities between them [2] [3]. This information can even be used to guide drug targeting decisions [4], namely how the targeting of one gene with a drug may reduce the risk and/or symptoms of a target disease while simultaneously inducing side effects because the gene is also associated with other traits [5] [6] [7]. On the other hand, genes associated with known disease risk factors can also be candidate drug targets [8]. A notable example of this is the targeting of low-density lipoprotein by inhibiting *PCSK9* expression to reduce coronary artery disease risk [9] [10]. The first step in inferring shared gene associations between multiple traits is generally to separately test each gene for association with each trait and to make a joint inference for the pair.

Inference of a shared gene association with multiple traits can be accomplished using standard hypothesis testing with gene-based test statistics from genome-wide association studies (GWAS) [11]. Then, a joint inference can be made using the intersection-union test at the SNP or gene level [12]. However, this process is highly sensitive to GWAS sample size, inherently limits the available interpretation only to nonzero disease association, and cannot be used to reliably characterize polygenicity genome-wide. Researchers also cannot make inferences about non-association at the gene level using hypothesis testing, which is required for identifying genes whose drugs have limited off-target potential. Hypothesis testing also relies almost entirely on the statistical power available to detect disease-associated SNPs in GWAS, which is primarily driven by the polygenic structure itself and GWAS sample size [13] [14]. For example, counts of rejected independent gene-based null hypotheses estimate the number of associated genes that exist for a single trait, but this estimate is biased in finite GWAS samples. Similarly, inference of no association between a gene and trait can only be validly made from a non-rejected null hypothesis when GWAS sample size is infinitely large.

These limitations make polygenic inference using hypothesis testing alone potentially unreliable and unflexible [15]. Instead, directly modelling the polygenic structure of complex traits and leveraging statistical methods which can supply a model-based posterior inference each trait in a pair may provide more consistent estimates of shared polygenicity and make a wider range of inferences available to researchers. However, currently available methods do not completely account for linkage disequilibrium correlation between their measured units (i.e., SNPs, genes) and can only provide polygenic inferences at the genome-wide or SNP levels [16] [17] [18], which in many applications are of at least secondary interest to the gene level.

We present a general statistical model which computes the posterior probability that each gene contributes to additive heritability using gene-based test statistics and an empirically derived model of polygenicity. This approach is leveraged to estimate the total number of genes which are putatively associated with each of 32 complex traits and the proportions of genes contributing shared and non-shared additive heritability across them. Our results suggest large variability across traits in the estimated number of associated genes, from less than 20 to over 900, and that the vast majority of disease associated genes are associated with multiple traits. These results highlight the role of prioritizing trait-exclusive risk genes as putative drug target candidates with less off-target effects. We show as an example that targeting the *KIT* gene for ALS prevention may reduce ALS risk while simultaneously increasing triglyceride levels, which could harm overall health. On the other hand, genes with evidence of pleiotropy across multiple traits can be leveraged if their associations are in the direction of risk, and we provide two demonstrative examples of this scenario for AD with *PLEKHA1* and ALS with *GAK*.

## Results

### Overview

We presented a <u>B</u>ayesian method to infer the <u>p</u>olygenic <u>a</u>rchitectures of <u>c</u>omplex traits (BPACT) under a model for SNP heritability of the *t*th complex trait using gene-based association test statistics from the set of all tested genes genome-wide 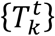. In this model, we estimate the proportion of causal genes 1 − *δ* and the outcome variance-scaled SNP heritability *τ*_*t*_ and calculate the posterior probability that the *k*th gene is associated with risk of the *t*th trait (PRP_*kt*_) as

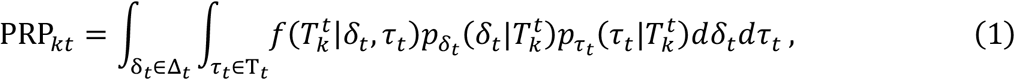

which integrates over the estimation error of (*δ*_*t*_, *τ*_*t*_) using the distributions 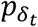 and 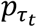 which are developed empirically. Integration over the spaces of the causal model parameters (*τ*_*t*_, *δ*_*t*_) allows us to account for misspecification of the model by its parameters. We infer the shared polygenic architecture for two traits *t* and *t*^*′*^ using {PRP_*kt*_} and {PRP_*kt*_^*′*^} while correcting for participant overlap between the GWAS from which 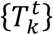 and 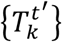 were calculated (*cf*. **Methods**). Inferences of genes with potentially low side effect potential are those for which

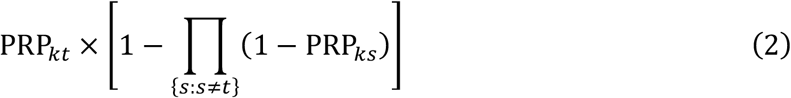

approaches 1. Inferences of potentially repurposable genes for the *t*th trait are those for which

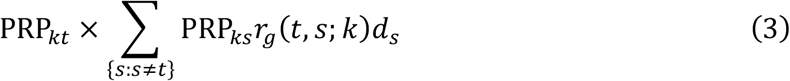

is high, where *r*_*g*_(*t, s*; *k*)*d*_*s*_ is the genetic correlation between traits *t* and *s* in the local region of the *k*th gene, i.e., *d*_*s*_ ∈ {1, −1}. We use the risk-directed genetic correlation *r*_*g*_(*t, s*; *k*)*d*_*s*_ to infer that the therapeutic targeting of the *k*th gene for the *t*th trait may be accomplished by targeting the *s*th trait. Although the PRP_*kt*_ quantity is a probability derived from a causal model, we limit our epidemiological inferences in specific disease contexts just to disease association and not causality, which could be better assessed using experimental approaches. We account for GWAS sample overlap between trait pairs in all analyses using a modified simulation extrapolation (SIMEX) approach [19] (*cf*. **Methods**). Results presented in the **Supplement** show that this removes any bias in estimated shared polygenicity between trait pairs, which for all trait pairs was small to begin with.

### Estimating polygenicity among 32 human complex diseases

We estimated the number of disease-associated genes (DAGs) for 32 complex traits and display the counts in **Figure 1a**. These results suggest that traits such as body mass index (BMI), high-/low-density lipoprotein (HDL/LDL), schizophrenia (SCZ), intelligence (INT), and diastolic, systolic, and pulse pressure (DBP, SBP, PP) may have more than 500 DAGs, while other traits such as Alzheimer’s disease (AD) or amyotrophic lateral sclerosis (ALS) may only have only 100-300. These counts are also highly correlated with the number of independent Bonferroni- and FDR-significant genes scaled by the square root of GWAS sample size, denoted respectively as sBonf and sFDR, which in the **Supplement** are shown to be proportional to the true number of disease associated genes. Hence, linear correlation of our estimated numbers of DAGs with sBonf (Pearson r=0.92) and sFDR (r=0.98) implies at least linear correlation between our estimates and the true numbers of true disease-associated genes. Independence between genes that were significant in hypothesis testing with gene-based association test statistics was inferred using the clumping procedure in [20] based on shared LD between gene-specific SNP sets. Our estimated counts of disease associated genes are also not correlated with GWAS sample size (Pearson r=0.02, P-value=0.900) or is square root (Pearson r=0.07, P-value=0.691) across the 32 traits, suggesting that they are not heavily influenced by statistical power in GWAS, the primary source of which is sample size.

**Figure 1:**
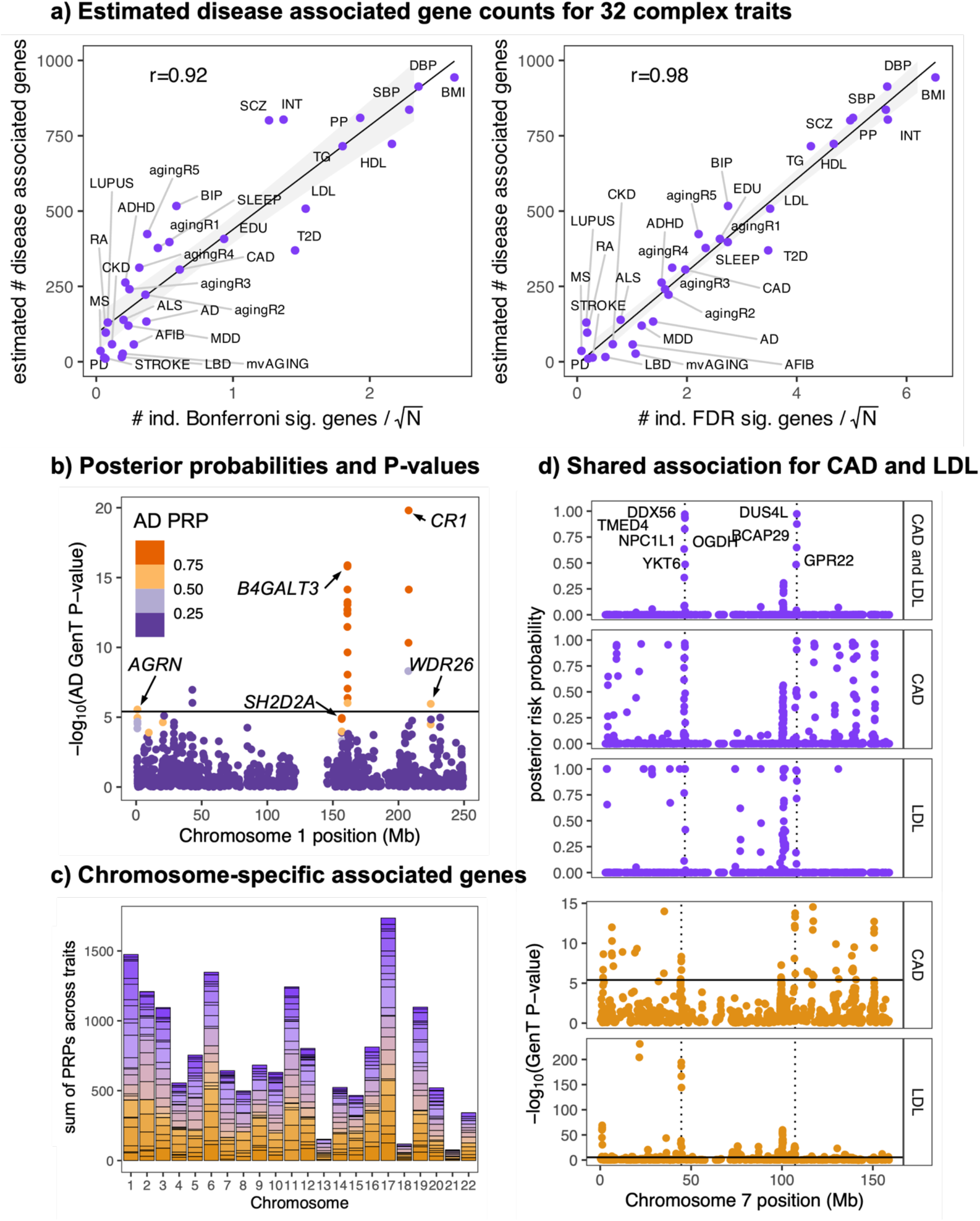
Estimated disease associated gene counts and example inference. **(a)**: Estimated counts of disease associated genes for 32 traits and their relationship with the number of Bonferroni (left) and FDR (right) significant genes using gene-based test statistics scaled by GWAS sample size. **(b)** Example of AD posterior risk probabilities (PRPs) for all tested genes on chromosome 1. **(c)** Estimated counts of disease associated genes on each chromosome from each of 32 traits calculated by using the prior distributions of *δ* (the empirically derived prior proportion of non-disease associated genes) for each trait. Different traits are represented by different colors which are separated by horizontal lines in each vertical bar. **(d)** Example of shared association on chromosome 7 for LDL and CAD. Probabilities in the ‘CAD and LDL’ panel are the products of LDL- and CAD-specific posterior risk probabilities and represent the posterior probability of shared association for each gene.

**Figure 1b** shows an example of the relationship between gene-level posterior risk probabilities (PRPs) using our method and gene-based association test P-values from the GenT method [20] for AD on chromosome 1. These results show that genes with the smallest P-values are most likely to be assigned a large PRP, such as *CR1* and *B4GALT3*. Though detecting gene-trait associations is not the primary goal of BPACT, genes which fail to meet the level of genome-wide significance in gene-based testing can still be assigned large PRPs under the BPACT model. One example was *SH2D2A*, which had PRP of 0.97 for AD but gene-based test P-value of 1.1E-5, above the Bonferroni significance level of 3.9E-6 (*cf*. **Methods**). *SH2D2A* is associated with neuronal signaling via synapse formation [21] and has previously been shown to be overexpressed in AD cases vs controls in immune cells [22], suggesting it may indeed be associated with AD risk.

**Figure 1c** displays the gene- and trait-specific posterior risk probabilities (PRPs) summed within each chromosome. These results suggest that the genes associated with the 32 traits are not uniformly distributed across the 22 chromosomes, but that chromosome 17 may contain the largest number of DAGs, despite it only containing the fifth largest number of tested genes (see **Supplementary Figure S9**). These counts are contributed from associated genes from all traits, and no small subset of traits dominates the chromosome-specific counts for any chromosome. We show in **Figure 1d** an example of how gene-level posterior risk probabilities can be used to provide an inference of shared association between coronary artery disease (CAD) and LDL, a known CAD risk factor [23]. These results highlight two lead loci on chromosome 7 which are likely shared between CAD and LDL, indexed by *DDX56* (7p13) and *DUS4L* (7q22.3), due to their posterior shared risk probabilities (S-PRPs) near 1.

### Estimating shared polygenicity among complex diseases

On average, an estimated 2.4 to 29.7% (mean=16.8%) of trait-associated genes are not shared with any other traits (**Figure 2a**). For example, of the estimated 120 genes which are inferred to associate with risk of major depression, only 14 (SE=0.89; 11.7%) are not associated with at least one other trait. Similarly, of the 58 genes inferred to associate with chronic kidney disease (CKD), only 13 (SE=0.81; 22.4%) are estimated to be specific only to CKD. We also estimate the total number of genes across the genome which are not associated with any of the 32 traits as 8,312 (SE=40.25), implying that approximately 50.9% (SE=2.5E-3) of the 16,324 genes tested for association with each of the 32 traits we studied may contribute to the SNP heritability of at least one of them (*cf*. **Methods**). **Figure 2b** shows the matrix of cosine similarity values [24] between all trait pairs, which is the ratio of shared disease associated gene counts to the geometric means of total disease associated gene counts for each trait pair. Larger cosine similarity values indicate greater degrees of associated gene sharing between trait pairs. The row/column ordering of traits in **Figure 2b** is determined by hierarchical clustering, and these results suggest that genetic similarity may be used to approximately group phenotypically similar traits into distinct clusters including a psychiatric/behavioral cluster, a metabolic/cardiovascular cluster, and an age-related/ autoimmune cluster. These clusters show greater evidence of associated gene sharing within them than between, and some traits such as stroke and chronic kidney disease show little evidence of gene sharing with any other trait, potentially explained by their low SNP heritability (**Supplementary Figure S4**). Together, these results suggest that cosine similarity indices applied to our estimates of shared association at the gene-level can be used to identify phenotypically similar traits, supporting our use of shared PRPs to identify shared genetic architecture between traits.

**Figure 2:**
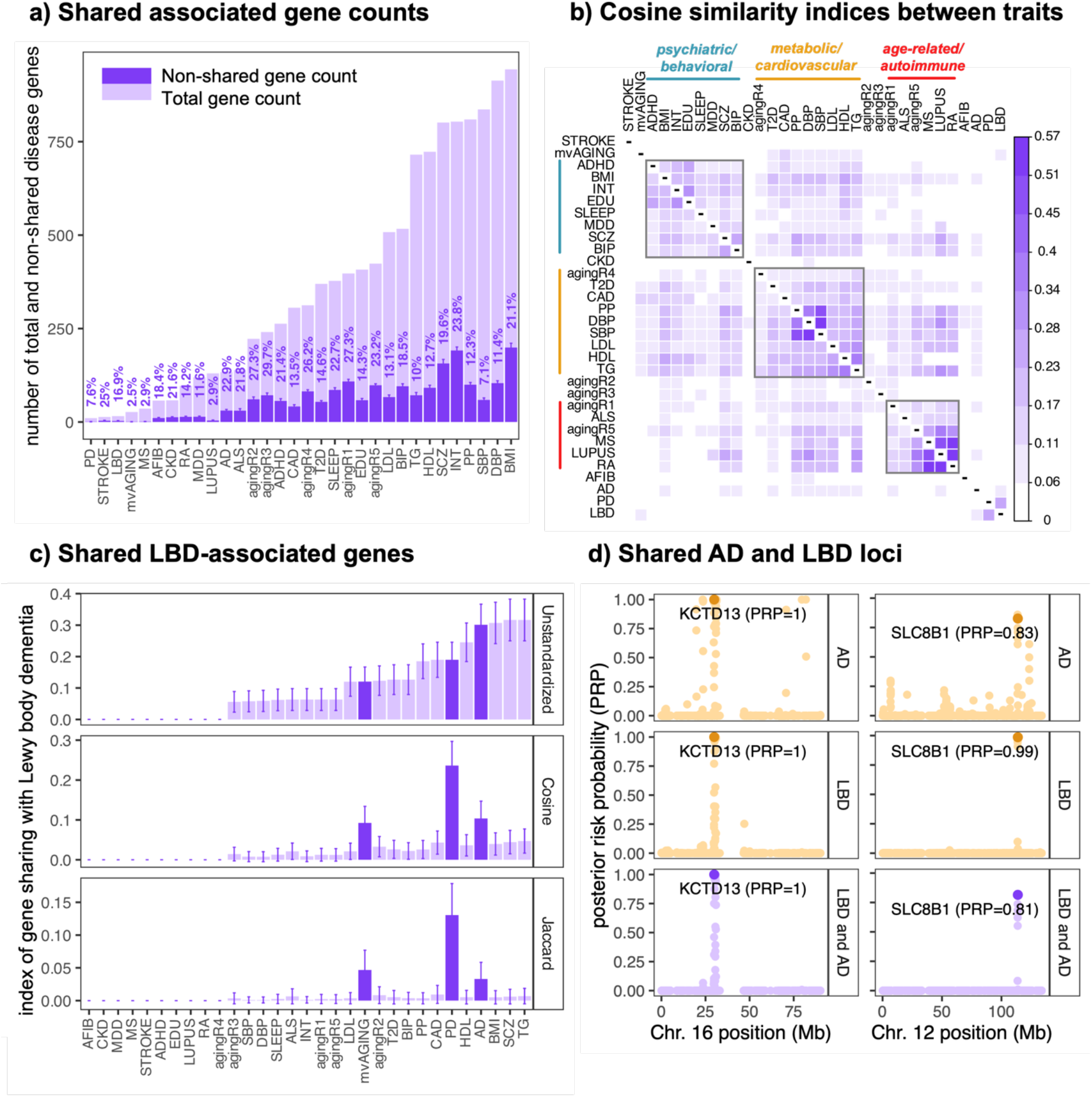
Measuring shared polygenicity across traits. **(a)**: Bars display estimated counts of all disease associated genes (light purple) and non-shared disease associated genes (dark purple). Vertically oriented percentages above each bar indicate the percentage of all disease associated genes that the number of non-shared genes represent. **(b)** Cosine index values between all pairs of traits (*cf*. Methods). Boxes are drawn around select sets of traits heuristically, and their labels assigned manually. **(c)** Estimated proportions of all LBD-associated genes which are shared with each other trait and their corresponding cosine and Jaccard index values (*cf*. Methods). **(d)** Examples of two loci with evidence of shared association for LBD and AD. ‘LBD and AD’ represents the posterior probability that each gene is associated with both LBD and AD risk.

A case study of polygenic sharing between traits is presented in **Figure 2c** for Lewy body dementia (LBD) using both the cosine and Jaccard [25] indices. These results show that the average LBD-associated gene is most likely to be shared with highly polygenic traits such as TG, SCZ, and BMI. But, after considering the estimated sizes of disease associated gene sets of LBD and the other traits, LBD is most genetically similar to Parkinson’s disease (PD), Alzheimer’s disease (AD), and a multivariate index of healthy aging (mvAGING; [26]). This is supported by their shared phenotype of age-related neurodegeneration. For example, there is an estimated 0.19 probability that a randomly selected LBD-associated gene is associated with PD risk, and an estimated 0.32 probability that a randomly selected LBD risk gene is associated with AD risk. We show in the **Supplement** that cosine index values for AD with the other 31 traits are mildly linearly correlated with their estimated genetic correlations (Pearson r=0.31) using LD score regression [13]. But, for some traits such as MDD, LDL, and EDU, there is evidence of a nonzero proportion of shared associated genes but no evidence of a nonzero global genetic correlation between them. Across all 496 trait pairs, the linear correlation between absolute genetic correlations and cosine index values was 0.22 (P=8.0E-7).

Two case studies of gene sharing between AD and LBD are presented in **Figure 2d** for two loci which are respectively indexed by *KCTD13* (16p11.2) and *SLC8B1* (12q24.13). *KCTD13* had a posterior probability of being shared by AD and LBD of 1.00 and is associated with brain morphology [27] and known to affect short-term memory by regulating synaptic activity in the hippocampus [28]. The inhibition of *SLC8B1* has also been shown to reduce cognitive performance and memory in mice [29] and its expression may protect against neuronal cell death [30]. Together, this supporting evidence suggests associations of *KCTD13* and *SLC8B1* with impaired memory, a hallmark symptom of both AD and LBD.

### Discovery of non-pleiotropic drug target candidates in ALS

We next provide a motivating example of how shared PRPs can be used to identify candidate drug targets for ALS with limited potential for off-target effects on other traits (*cf*. Equation 8). **Figure 3a** displays PRPs for ALS and for ALS but not any of the other 31 traits using the set of 3,369 genes with drug targets (i.e., ‘druggable genes’; *cf*. **Methods**). These results suggest that *TBK1* and *SCNN1B* have the largest PRPs (0.87 and 0.89, respectively) of associating with ALS but none of the other 31 phenotypes. This suggests they may have lower off-target potential compared to other drug targets with respect to side effects on any of the other 31 traits. As a counterexample, the *KIT* gene is associated with ALS, triglycerides (TG), high-density lipoprotein (HDL), Bipolar I/II disorder (BIP), and body mass index (BMI). **Figure 3b** shows the SNP-level associations between ALS, TG, and gene expression in the thyroid from GTEx v8 [31] in the *KIT* locus. These results suggest the presence of nonzero associations between SNPs in this locus and ALS, TG, and *KIT* expression. There is also evidence of a negative local genetic correlation between ALS and TG and between ALS and thyroid gene expression, and a positive local genetic correlation between TG and thyroid gene expression. Mendelian Randomization (MR) using all FDR-significant thyroid eQTLs as instruments and observed eQTL and ALS Z-statistics as in [32] suggested a negative causal effect of *KIT* expression on ALS risk (Estimate= –0.12; P=1.8E-8). These results suggest that while decreased *KIT* expression in the thyroid may reduce ALS risk, this may simultaneously be associated with increased TG levels, potentially harmful to overall health.

**Figure 3:**
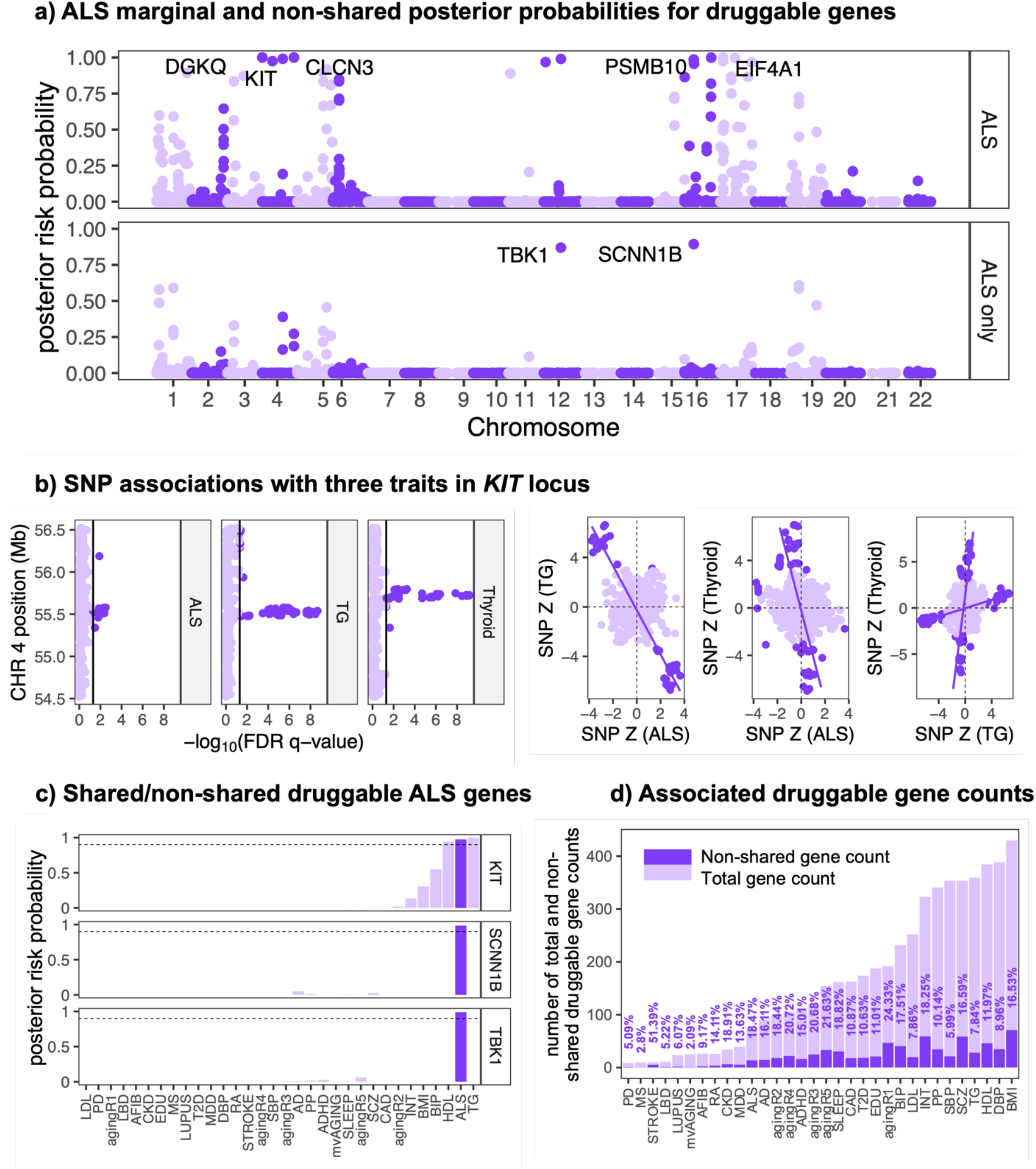
Shared association of ALS druggable genes with other traits. **(a)** Genome-wide plot of posterior risk probabilities (PRPs) for ALS (top) for each gene and posterior probabilities that each gene is associated with ALS but not with any of the other 31 studied traits (bottom). **(b)** (left) SNP-level association estimates from GWAS for ALS, TG, and gene expression in the in the *KIT* locus (Chr4: 54524085-56606881). (right) Bivariate SNP associations between ALS, TG, and thyroid eQTLs in the *KIT* locus. Dark purple points are FDR significant at the 5% level for either trait on the x- or y-axis. Lines are of best fit through sets of FDR-significant SNPs. **(c)** posterior risk probabilities of three select genes for association with each of the 32 studied traits. Vertical bars corresponding to ALS are colored in dark purple; bars not corresponding to ALS are colored in light purple. **(d)** Bars display estimated counts of all druggable disease associated genes (light purple) and non-shared druggable disease associated genes (dark purple). Vertically oriented percentages above each bar indicate the percentage of all druggable disease associated genes that the number of non-shared druggable disease associated genes represent.

The sharing of disease association between *KIT* with ALS and TG, and the corresponding non-sharing of the *SCNN1B* and *TBK1* genes with ALS only, is demonstrated in **Figure 3c**. We note that non-sharing of association between ALS and non-ALS traits is not a requirement for a putatively safer ALS drug target, but that the set of genes with low non-ALS trait associations may contain genes with less complex biological pathways of effect and hence better candidates as therapeutics with limited side effect potential. **Figure 3d** displays estimated counts of shared and non-shared disease associated genes that have drug-target interactions for all traits, made by directly summing PRPs for these genes. These results suggest that for some traits such as multiple sclerosis (MS), PD, and LBD, currently available drug targets are likely to associate with at least one other trait. These results also suggest that approximately 14 druggable genes are candidate ALS targets with limited evidence of conferring off-target effects by modifying any of the other 31 traits. Of these 14, the *TBK1* and *SCNN1B* genes have the strongest evidence of ALS association and limited non-ALS association. We provide trait-specific PRPs and pairwise trait-shared PRPs for all 3,369 druggable genes for all traits in the **Supplemental Data**.

### Discovery of non-pleiotropic drug target candidates in Alzheimer’s disease (AD)

We also leveraged estimated shared polygenicity between AD and each of the other 31 traits to identify candidate drug targets which have limited evidence of side effect potential. Some examples of these genes include *EARS2, EPHA1, ITGA2B, MME*, and *MS4A2*, each of which have a PRP with AD greater than 0.9, and no PRP with any other trait greater than 0.9 (**Supplementary Figure S10**). The *MS4A2* gene is a member of the *MS4A* gene cluster (11q12.1) which is known to play a critical role in autoimmune activation [33] [34] and even moderate effects of the well-known AD risk gene *TREM2* [35] [36]. On the other hand, many well-known druggable AD risk genes have evidence of pleiotropic association with other traits (**Supplementary Figure S10**). These genes include *ACE, APOC2, APOE, BIN1*, and *CLU*. As examples, *CLU* had PRP with AD of 0.99 and with SCZ of 1.00; *APOC2* had PRP with AD of 1.00 and PRPs with TG, LDL, HDL, and CAD each of 1.00. *CLU* may simultaneously confer risk of AD and SCZ because of its role in and response to autoimmunity and inflammation, which is dysregulated in both AD and SCZ cases vs controls [37] [38] [39]. *APOC2* is known to be involved in the metabolism of lipoprotein [40], and so has been implicated in hypertriglyceridemia risk [41], suggesting that its conferral of AD risk may be mediated by lipidemia.

### Repurposable drug candidates using BPACT

We leveraged the shared association of a gene with multiple disease trait(s) to identify candidate drug targets that may have a therapeutic effect on multiple diseases simultaneously. We refer to these genes as repurposable drug candidates since they may already have some demonstrated evidence of treating one condition but have so far not been extensively evaluated for the treatment of another. For these genes, a genetic correlation between the target disease and other traits in the direction of risk (e.g., positive correlation for T2D and LDL cholesterol, negative correlation for T2D and intelligence), suggests that targeting them with a drug may have therapeutic effects on each trait simultaneously. We identified these genes as those with large PRPs for multiple traits with which the index trait was locally genetically correlated in the direction of risk with all other traits on average (*cf*. Equation 3) and present these quantities in **Figure 4a**. These results suggest that traits such as SCZ, CAD, and T2D may have the largest number of repurposable candidates, which is expected since these traits have evidence of being highly polygenic (**Figure 1a**), and that traits such as lupus and MS may not have any known associated genes that are not associated with at least one other trait.

**Figure 4:**
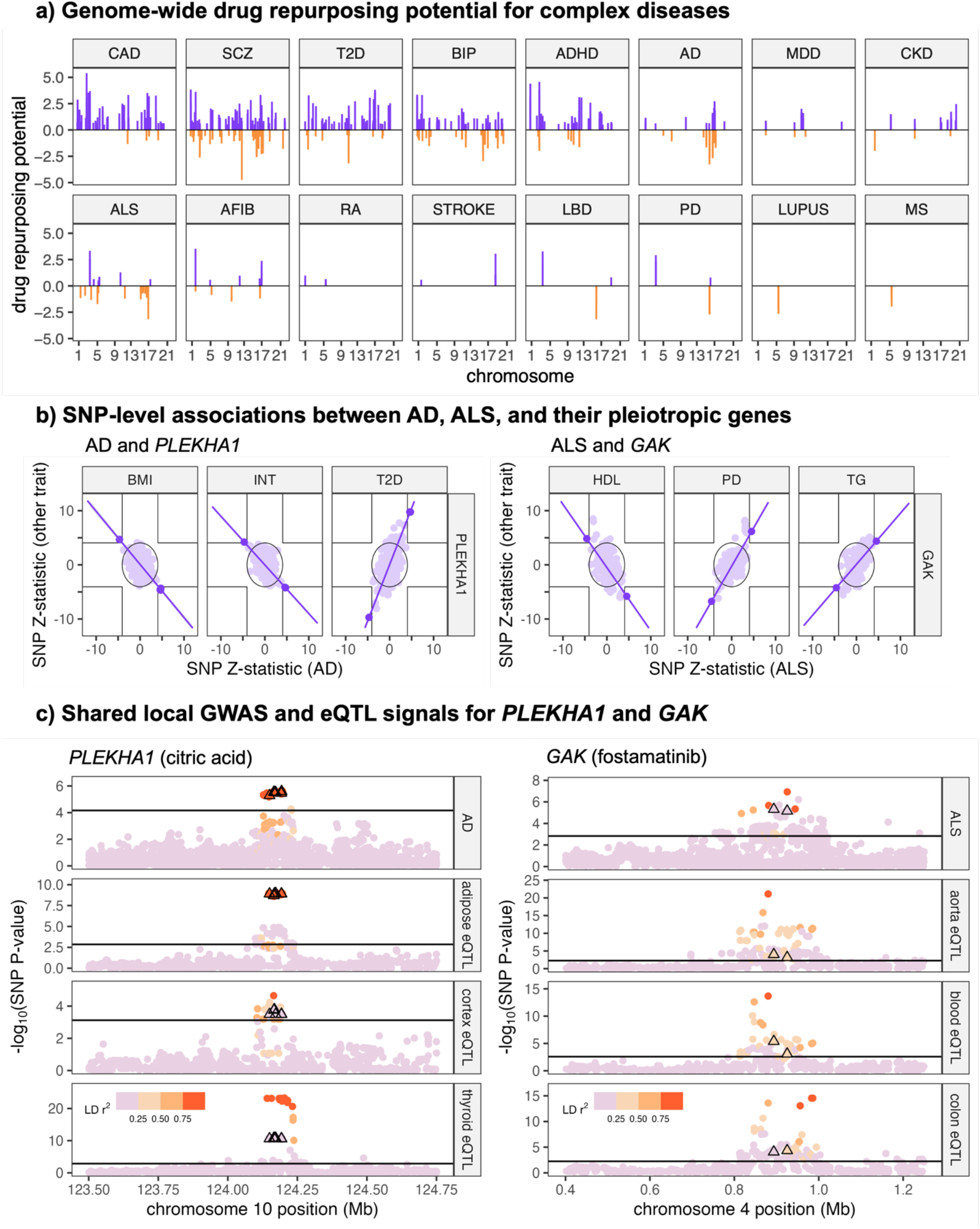
Drug repurposing targets for disease traits. **(a)** Drug repurposing potential values (*cf*. Equation 9) for the 16 complex diseases we studied and each tested druggable gene. Positive values denoted by purple bars are candidate repurposable targets. **(b)** Examples of candidate repurposable drug targets for AD with *PLEKHA1* (left) and ALS with *GAK* (right). Displayed are Z-statistics for association at the SNP level from each respective GWAS for the indicated traits in the ±100Kb window around each gene body in hg19 coordinates. Dark purple points are significant at P<5E-5 in an intersection union test for the pair of traits on the x- and y-axes. Dark purple lines correspond to the best linear fit through these points. The null region of the IUT at the 5E-5 level is indicated by the interior of joined vertical and horizontal lines; the null region of a joint test at the 5E-5 level is indicated by the interior of the circle centered at the origin. **(c)** Local SNP associations between *PLEKHA1*, AD, and select tissue contexts of gene expression (left) and between *GAK*, ALS, and select tissue contexts of gene expression (right). The color of each point represents its squared LD with the lead SNP, which was estimated using the 1KGv3-EUR reference panel (Siva et al., 2008). SNPs represented by triangles correspond to the SNPs which are highlighted in dark purple in panel **(b)**.

As a demonstrative example, we show in **Figure 4b** that *PLEKHA1* is associated with BMI, intelligence (INT), and Type 2 diabetes (T2D), and that the directions of their local genetic correlations with AD suggests that targeting *PLEKHA1* may have therapeutic/preventative effects on all traits including AD. In this locus, six SNPs (rs10788284, rs6585827, rs2421016, rs7097701, rs10510110, rs2280141) are associated with both AD and each of BMI, INT, and T2D at level P<5E-5 in intersection-union tests (IUTs). The rs10510110 SNP of *PLEKHA1* was associated with AD (FDR=3.2E-3) and gene expression (GTEx v8; [31]) in visceral adipose (FDR=3.6E-7), brain cortex (FDR=3.4E-2), and thyroid tissue (FDR=1.8E-9) (**Figure 4c**). Citric acid (CA) is a nutraceutical agent interacting with *PLEKHA1* (DrugBank, DB04272; [42]) that is traditionally recognized for its association with BMI, lipid cholesterol, and glucose metabolism [43] [44]). There is additional evidence suggesting that CA can modulate oxidative stress in the brain by reducing inflammation and lipid peroxidation [45], potentially via its role in fatty acid metabolism [46] and/or its inhibition of the acetylcholinesterase (AChE) enzyme [47]. AChE inhibitors have demonstrated efficacy in treating AD symptoms [48], and one hypothesized mechanism is via the deformation of amyloid-beta tangles [49] [50].

Similarly to AD and *PLEKHA1*, two SNPs in the *GAK* locus (rs3775121, rs873785) were associated with HDL, PD, and TG (P<5E-5), and their local genetic correlations with ALS were each in the direction of risk for all traits. The rs3775121 SNP was associated with ALS (FDR=5.3E-3) and gene expression (GTEx v8; Lionsdale et al., 2018) in aortic (FDR=1.4E-3), blood (FDR=2.8E-4), and colon tissue (FDR=2.2E-3) (**Figure 4c**). Fostamatinib is a drug which primarily inhibits spleen-associated kinase (*SYK*), but can also inhibit many additional kinases including cyclin-G-associated kinase (*GAK*) (DrugBank, DB12010; [42]). Fostamatinib was originally developed to treat immune conditions such including autoimmune hemolytic anemia, thrombocytopenia, and immunoglobulin A nephropathy [51], though bioinformatic analyses using MR and molecular docking support fostamatinib as a potential therapeutic target for ALS and PD [52] [53] [54]. Fostamatinib may have evidence as a candidate ALS target because *SYK* and *GAK* can cause inflammation-associated cell death and cognitive impairment [55] [56] [57] [58], a hallmark symptom of ALS.

In summary, BPACT identified genes which may be drug repurposing candidates for complex traits including ALS and AD. These genes had evidence of association with multiple traits each in the direction of risk such that targeting them with a drug could lead to therapeutic effects for all associated traits. We demonstrated this phenomenon using the AD-associated *PLEKHA1* gene and ALS-associated *GAK* gene. These genes interact with available compounds – citric acid with *PLEKHA1* and fostamatinib with *GAK* – and the literature provides supporting evidence that these compounds associate with AD and ALS pathology. Future functional and experiment studies may provide greater insight into the degree to which these compounds may simultaneously confer protective effects against AD/ALS and other traits.

## Discussion

We present BPACT, which can be used to estimate the number of genes which contribute to the SNP heritability of a single trait or multiple traits. We used BPACT to show that more than half of the genome has evidence of explaining SNP heritability in at least one of the 32 traits we studied. Results from analyses of shared heritability highlighted druggable genes that have evidence of reducing disease risk at the potential cost of side effects, and we demonstrated this phenomenon using *KIT* for ALS as an example. Our results also showed how BPACT can be used to identify genes which are candidate therapeutic targets for multiple traits simultaneously, and we demonstrated this used AD and ALS as examples.

We began by showing that there is substantial variability in the estimated number of disease associated genes (DAGs) across the 32 traits. BPACT was also able to identify a highly polygenic group of traits which included BMI, DBP, SBP, pulse pressure (PP), LDL, HDL, TG, INT, BIP, and SCZ. Estimates of polygenicity correlated almost perfectly with the number of GWAS significant genes adjusted for sample size, which we showed in the **Supplement** to be proportional to the true number of associated genes for a given trait. This concordance provides supporting evidence that our estimator of the number of disease associated genes is at least proportional to the true number of disease associated genes for the trait. Since this quantity is adjusted for GWAS sample size, it may not be strongly related to GWAS statistical power. We then showed that the estimated number of disease associated genes (DAGs) across all 32 traits is not uniformly distributed across chromosomes after adjusting for the number of genes they contain (**Supplement Figure S9**), and that chromosome 17 had the largest estimated number of genes which were putatively associated with any trait.

We next showed that most genes which explained nonzero heritability in a trait were shared with at least one other trait. For example, we estimated that 24.9% of genes associated with ischemic stroke risk and only 2.5% of genes associated with the index of aging (mvAGING) were not associated with any other trait we tested. We then showed that cosine and Jaccard indices applied to total and shared DAG counts can be used to measure their shared polygenicity. These quantities were reported in the **Supplement** to moderately positively correlate with estimated global absolute genetic correlations. But, for some trait pairs, we find evidence of substantial sharing but estimated genetic correlations very close to 0. These results provide empirical evidence that shared polygenicity is a weaker form of genetic similarity than genetic correlation, and that global null estimates of genetic correlation do not imply genetic independence between traits.

The evidence also suggested that most trait-associated genes with known drug interactions were shared across multiple traits, and we showed an example of the *KIT* gene associated with ALS, BIP, TG, HDL, and BMI. Expression of *KIT* in the thyroid was negatively associated with ALS risk but positively associated with TG levels, suggesting that targeting of *KIT* with an inhibitor may reduce ALS risk but simultaneously elevate TG levels, a potentially harmful side effect. We presented the *TBK1* and *SCNN1B* genes as examples of alternative druggable candidates, each of which were associated with ALS but had no evidence of association with any of the other 31 traits. *TBK1* can mediate activation of the NF-*κ*B transcription factor which regulates innate and adaptative immunity processes [59] [60] [61], and targeting of *TBK1* with fostamatinib has been demonstrated to reduce ALS risk in vitro [53]. *SCNN1B* has been shown to suppress the MAPK signaling pathway [62] which is dysregulated in ALS patients [63] [64].

Our results also showed that leveraging gene pleiotropy across multiple traits may nominate repurposable drug targets, and two examples included *PLEKHA1* with AD and *GAK* with ALS. These targets can be viewed as potentially therapeutic for an index trait because of modification to one or more of its risk factors, or simultaneous therapeutic effects on multiple traits via independent biological pathways. *PLEKHA1* is expressed across many tissues including the brain, heart, and adipose tissue (GTEx v8; [31]), and contains the pleckstrin homology protein folding domain which is associated with the binding of phosphorylated lipids onto inositol rings [65]. Knockout of *GAK* has been shown to disrupt the homeostasis of lysosomes during autophagy [57], and like *PLEKHA1* it is expressed across many tissues including the brain, heart, and adipose tissue [31]. The pleiotropic effects of these genes on multiple phenotypically distinct traits may be explained by their broad expression patterns across multiple tissues. We showed that the proportion of genes for which shared associations with other traits can be detected is generally large, emphasizing that the richest source of candidate drug targets for a complex disease may be provided by the set of pleiotropic genes.

Our study is strengthened by the generality of the underlying statistical model and the wide range of inferences which can be made once it is fitted. The model is also advantageous because it accounts for its own parameter misspecification by integrating over the prior parameter space. Gene-level posterior risk probabilities can be used to infer trait-specific association, trait-shared association, and trait non-association in the context of the BPACT statistical model. We also provide expressions for the effect of GWAS sample overlap on gene-based test statistic correlations and the working number of independent gene-based association tests genome-wide (*cf*. **Methods**). These quantities have henceforth been absent in the literature and have respectively precluded direct comparison of gene-based test statistics from multiple GWAS cohorts and well-calibrated control of Type I error rates in genome-wide testing. We proposed a new approach to address the challenge of correlated gene-based test statistics (*cf*. **Methods**) during inference, which is based on the well-studied SIMEX approach [19]. We show in the **Supplement** that this procedure removes any extant bias in estimated counts of shared associated genes between trait pairs due to GWAS sample overlap. We also present a new model-fitting approach for correlated gene-based test statistics based on composite posterior densities which are iteratively evaluated over randomly selected and weakly correlated gene sets (*cf*. **Methods**). This approach uses the principles of imputation to make its inference, and without it our approach could lead to inflated estimates of disease associated gene counts because of shared LD between gene-specific SNP sets. Finally, **Supplementary Table S1** shows that our method is computationally efficient, spending approximately 15 minutes to run on an Intel® Xeon® Gold 6148 CPU 2.40GHz machine.

BPACT has the following limitations. Estimates of DAG counts and their shared proportions adjust for GWAS sample size, but do assume accurate phenotyping, correct specification of the gene-based test statistic null distributions, and that genes only have non-interacting effects on the trait. Violations of any of these assumptions may bias the estimated DAG counts via mis-specified model priors and likelihood functions. Our model intends to account for misspecification of model parameters, but does assume a correctly specified model structure of additive SNP heritability. If the structure and/or parameters of these models are inappropriate for some traits, or for some loci for some traits, it may cause the model to return a gene-level PRP which is far from the true value of the latent binary indicator of association. This could have downstream negative consequences on estimated shared and non-shared DAG counts for traits and their pairs. Our model also assumes that within each block of correlated gene-based test statistics, only a single disease associated gene is present. Future extensions of the BPACT method may attempt to relax this assumption. Finally, an inherent limitation of using gene-based test statistics to make the aforementioned inferences is that any nuance at the SNP level may not be completely captured at the gene level. However, this aggregation across SNPs in a gene-specific set is what makes gene-based test statistics statistically powerful [20].

In addition to inferences related to drug target identification and polygenicity evaluations, future researchers may also use BPACT and its results to evaluate the degree to which increases in GWAS sample sizes are likely to detect additional genes explaining meaningful amounts of heritability. For example, we estimate that 133 genes associate with AD, and a recent AD GWAS has detected 82 independent loci [66]. This suggests that increasing AD GWAS sample size may be likely to produce new meaningful insights into the genetic etiology of AD. However, for traits such as BMI, for which we estimate that approximately 943 associated genes may exist, the currently reported ∼1,000 BMI-associated loci [67] may cover most of the entire associated gene set and so further investment in continually larger BMI GWAS sample sizes may not return the same value which it requires. Finally, the integration of quantitative trait loci (QTL) data into the BPACT model may help it to nominate drug targets with supporting transcriptomic, proteomic, epigenomic, and/or metabolomic evidence that may have greater therapeutic potential than those identified just from GWAS summary statistics.

## Methods

### SNP-level model

Our gene-level model is built from a SNP-level random effect model for a single trait. Let 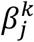 represent the marginal association between the *j* th SNP of the *k* th gene and a trait, *r*_*jj*_^*′*^ the LD correlation between SNPs *j* and *j*^*′*^ corresponding to the gene, *𝓁*_*j*_ the LD score of the *j* th SNP (i.e., sum of squared LD correlations with surrounding SNPs in a window of fixed size; [13], *a*_*j*_ the corresponding minor allele frequency, *N* the GWAS sample size, and 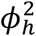 the average SNP heritability explained by each of 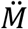 causal SNPs genome-wide. Let *G*_*ij*_ represent the dosage of risk alleles of the *j* th SNP from the *i*th person in the GWAS and 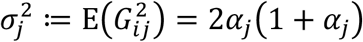. We use the following SNP-level models:

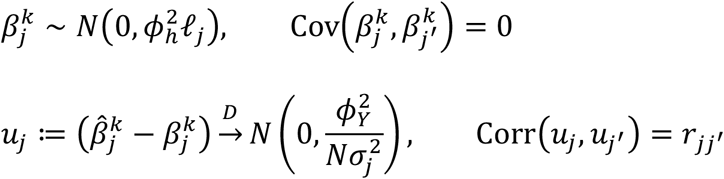

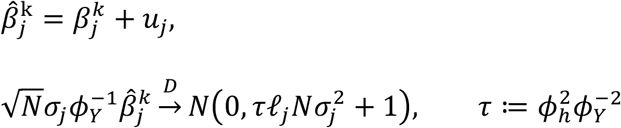

where 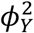 is the scaled conditional variance of the phenotype given the genotype for the *j*th SNP. For binary phenotypes, 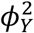 acts also as a scaling factor which is proportional to the trait prevalence that in effect indicates a transformation from the observed binary scale to the underlying liability scale [68]. It follows from the definition of *τ* that 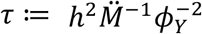 is likely to be very small in practice when 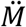 is large and/or 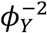 is large and SNP heritability *h*^2^ is small. Let 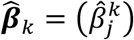 be the vector of estimated SNP effect sizes for all SNPs in the set corresponding to the *k* th gene. The correlation matrix of 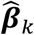 is approximately the LD matrix **R**_*k*_ under this model [69].

### Gene-level model

The SNP-level model is used to construct a gene-level model for the distribution of gene-based test statistics {*T*_*k*_}. Let the *k*th of *M* genes across the genome be tested using the statistic *T*_*k*_ calculated from *m*_*k*_ SNPs in the set *𝒮*_*k*_, i.e.

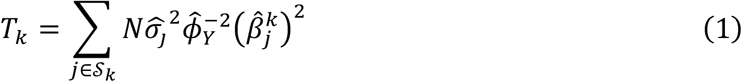

[20] showed that under *H*_0*k*_: E(*T*_*k*_) = *m*_*k*_, *T*_*k*_ *∼* Γ(*α*_0*k*_, *ξ*_0*k*_) approximately and under *H*_1*k*_: E(*T*_*k*_) < *m*_*k*_, *T*_*k*_ *∼* Γ(*α*_1*k*_, *ξ*_1*k*_) approximately, using the shape-rate parameterizations of each Gamma distribution. Let 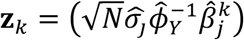, **L**_*k*_ = diag(*𝓁*_*j*_) and 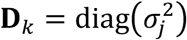 for *j* ∈ *𝒮*_*k*_, and 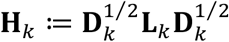 such that Cov(**z**_*k*_|*H*_1*k*_) = *Nτ***H**_*k*_ + **R**_*k*_ and Cov(**z**_*k*_|*H*_0*k*_) = **R**_*k*_. It is shown in the **Supplement** that

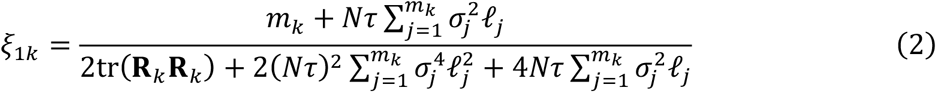

and

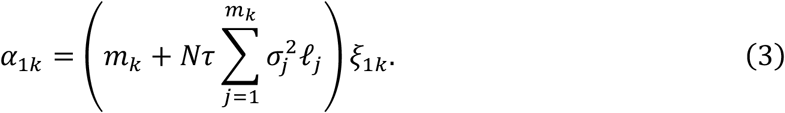

It follows immediately that *ξ*_0*k*_ = *m*_*k*_tr(2**R**_*k*_**R**_*k*_)^}1^ and *α*_0*k*_ = *m*_*k*_*ξ*_0*k*_. We use these quantities to model the data-generating process of the statistic *T*_*k*_ as

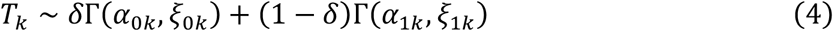

where 1 − *δ* is the marginal probability that gene *k* is causally associated with the phenotype under this model. This mixture can alternatively be expressed as

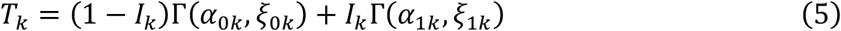

where P(*I*_*k*_ = 1) = 1 − *δ*, P(*I*_*k*_ = 0) = *δ*, and *I*_*k*_ is a latent indicator of causality for the *k* th gene.

### Estimating the number of disease associated genes and their shared counts

We presented a <u>B</u>ayesian method to infer the <u>p</u>olygenic <u>a</u>rchitectures of <u>c</u>omplex traits (BPACT) and the sharing of architectures between pairs of them. In this model, we calculate the posterior probability that the *k*th gene is associated with risk of the *t*th trait (PRP_*kt*_) as

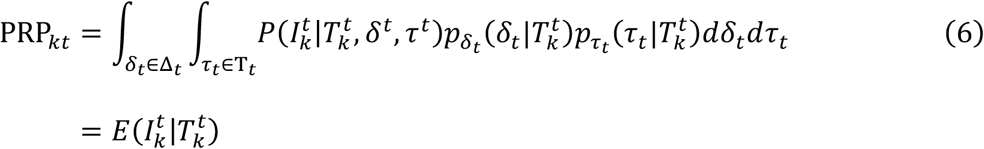

where 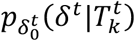 and 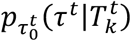 are posterior distributions of *δ*^*t*^ and *τ*^*t*^, respectively, which are assumed conditionally independent given 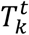, and

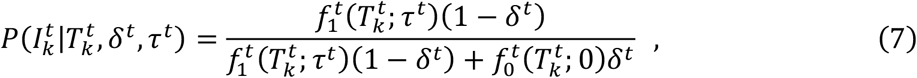

where 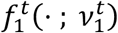 and 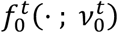 respectively are the non-null and null distributions of 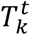 parameterized by 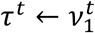 and 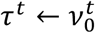. The quantity PRP_*kt*_ is interpreted as the posterior probability that gene *k* causes trait *t* given the statistic 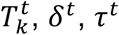, *δ*^*t*^, *τ*^*t*^, and their distributions 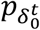 and 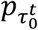 in the spaces Δ_*t*_ and T_*t*_, which can be interpreted as adjustments for the estimation error of 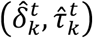. We estimate the posterior distributions 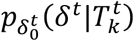 and 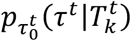 by sampling from their distributions computationally using the Metropolis-Hastings algorithm described in the **Supplement** and thereafter model them with Beta and Trapezoidal distributions, respectively.

We infer the shared polygenic architecture for two traits *t* and *t*^*′*^ using {PRP_*kt*_} and {PRP_*kt*_^*′*^} while correcting for participant overlap between the GWAS from which 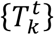 and 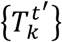 were calculated (see the next subsection). Inferences of genes with potentially low off-target effects are those for which

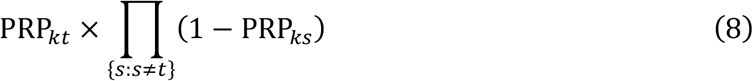

is high, and inferences of potentially repurposable genes for the *t* th trait are made for those for which

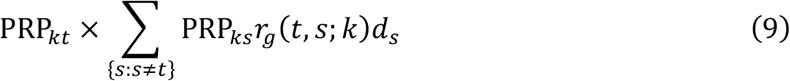

is high, where *r*_*g*_(*t, s*; *k*)*d*_*s*_ is the re-signed genetic correlation between traits *t* and *s* in the local region of the *k*th gene, i.e., *d*_*s*_ ∈ {1, −1}. We use the risk-directed genetic correlation *r*_*g*_(*t, s*; *k*)*d*_*s*_ to infer that the therapeutic targeting of the *k*th gene for the *t*th trait may be accomplished by targeting the *s* th trait. Although PRP_*kt*_ is a conditional probability under a model representing statistical causality, we describe our practical epidemiological inferences in specific disease contexts just to disease risk association and not causality, which could be better assessed using experimental approaches.

We estimate the total number of disease associated genes for trait *t* using *S* randomly selected, mutually weakly correlated, and chromosome-specific sets of genes, denoted as *𝒞*_*r*_(*s*) for chromosome *r* = 1, …, *R* at the *s*th iteration, and pooling results across the *S* replicates and *R* chromosomes to produce the estimate:

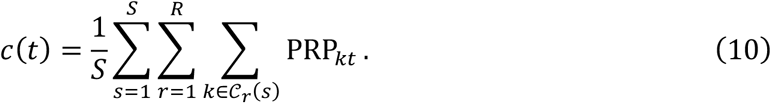

We can estimate the number of shared associated genes between traits *t* and *t*^*′*^ as

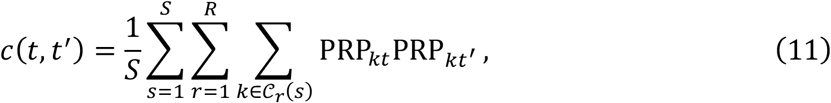

which assumes independence between 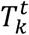 and 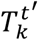 for all *k*. Independent sets of genes from each chromosome at the *s*th iteration are constructed such that the correlations between all pairs of gene-based test statistics in the set *𝒞*_*r*_(*s*) are below a fixed level, which in practice we set as √0.5. Since many of such sets may exist, we first define approximately independent blocks of genes using the √0.5 threshold for Corr(*T*_*k*_, *T*_*kt*_) and the correlation block-sorting method of [70] in the bigsnpr R package. For each block and chromosome at each iteration, we randomly select one gene and add it to the set *𝒞*_*r*_(*s*). Across the 16,324 genes which were tested for association with all 32 traits in our real data analysis, 8,148 independent blocks of gene-based test statistics were present. The inferences of total, *c*(*t*), and shared, *c*(*t, t*^*′*^), association are therefore based on composite posterior densities fitted multiple times for *S* iterations, over which the results are pooled to produce the final estimates. We calculate standard errors for estimated quantities using the imputation principles of [71] across the *S* iterations. We estimated the number of genes which are associated with any trait by first subtracting from 1 the product of all trait-specific posterior risk probabilities and summing them across all independent genes. When then divided this quantity by the estimated number of independent gene blocks, 8,148, and multiplied it by 20K.

### Effect of sample overlap on estimated shared gene counts

In the previous subsection, we introduced *c*(*t, t*^*′*^) which assumed 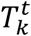 was uncorrelated with 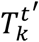 for the *k*th gene and *t*th, *t*^*′*^th phenotypes. We show in this subsection that overlapping subjects between the GWAS in which 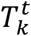 and 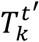 were calculated can induce nonzero spurious correlation between 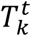 and 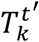, and how we can correct for it. For generality and to illustrate the motivation for considering independent blocks of genes in our polygenicity estimation, we show the correlation between gene-based test statistics in the case of different genes and for two traits from different GWAS cohorts which may contain overlapping subjects. Let **R**_*kk*_^*′*^represent the matrix of LD correlations between SNPs in the gene-specific sets for genes *k* and *k*^*′*^, ***𝚼*** _*tt*′_ = (*υ* _*tt*′_), and *υ* _*tt*′_ ≈ *N*_*tt*′_ (*N*_*t*_*N*_*t*′_)^−8/2^Corr(*t, t*^*′*^) [72], which represents the approximate correlation between 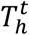 and 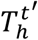 for any gene index *h* using GWAS of continuous traits *t* and *t*^*′*^ of sizes *N*_*t*_ and *N*_*t*′_ containing *N*_*tt*′_ overlapping subjects. For binary traits, *υ* _*tt*′_ will have a slightly different expression but the general principle is the same [72]. It follows that

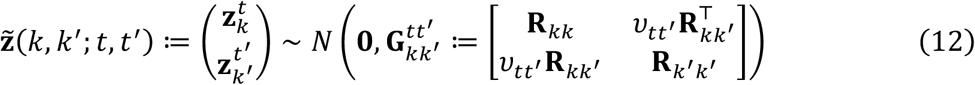

under 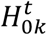 and 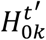 and that 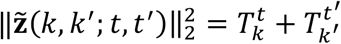. It remains to find 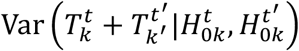, which is equal to 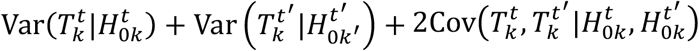 and where

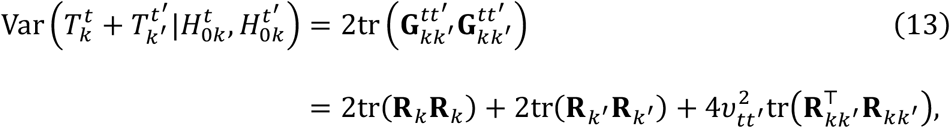

since 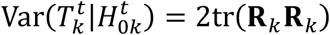 and 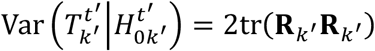, implying that

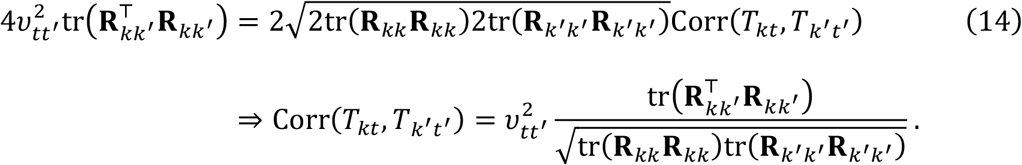

This quantity is strictly positive and states that the correlation between gene-based test statistics from different genes and for different traits from separate GWAS is proportional to the product of shared LD between gene-specific SNP sets, the proportion of shared GWAS subjects, and the phenotypic correlation between the traits. The quantity 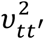 has as its maximum the square of phenotypic correlation Corr(*t, t*^*′*^) in a single GWAS cohort. This shows that any pair of gene-based tests statistics, and by extension PRP_*kt*_ and PRP_*kt*′_, are not generally independent if there are overlapping GWAS subjects and SNPs in LD between the SNP sets. This motivates correction of *c*(*t, t*^*′*^) for nonzero correlation between PRP_*kt*_ and PRP_*kt*′_.

To perform this correction in practice, researchers can either (i) de-correlate SNP-level Z-statistics for each trait pair using the method of [72] before applying gene-based association testing and subsequent polygenicity evaluations to each trait, or (ii) apply our post-hoc correction to the estimated number of shared disease associated genes using the principles of simulation extrapolation (SIMEX; [19]). We intend to measure the effect of GWAS sample overlap on estimates of shared disease associated gene counts between pairs of traits using simulation because an analytic expression of it using the definition of *c*(*t, t*^*′*^) is challenging to derive. In this procedure, we begin by estimating posterior risk probabilities PRP_*kt*_ and PRP_*kt*′_ for traits *t* and *t*^*′*^ and all genes *k* = 1, …, |*𝒮*(*t, t*^*′*^)| in the set of genes *𝒮*(*t, t*^*′*^) tested for association with both traits. We require the estimated number of disease associated genes *c*(*t*) and *c*(*t*^*′*^) for each trait, their estimated SNP heritability or their *τ*^*t*^ and 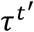 values, and the estimated sample overlap correlation parameter *υ* _*tt*′_ for the trait pair, which is estimable from GWAS summary statistics as the empirical correlation between non-significant SNP-level Z-statistics [32].

We then specify a grid of 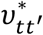 values in the interval 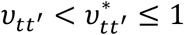, generate simulated SNP-level summary statistics under the above model which includes GWAS sample overlap, perform gene-based association testing using the sum of SNP chi-squares, estimate gene-level posterior risk probabilities for each trait directly from the likelihoods of the latent indicators (i.e., without integrating over the prior distributions of *δ* and *τ*), and estimate the number of shared disease associated genes using the *c*(*t, t*^*′*^) estimator with |*𝒮*(*t, t*^*′*^)| = 8,148 independent genes. We use the simulation-averaged estimates of shared counts at each 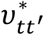 value to extrapolate from the observed shared count *c*(*t, t*^*′*^) at *υ* _*tt*′_back to the estimated shared count when there is no GWAS sample overlap, denoted as 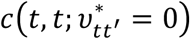. We then multiply the original *c*(*t, t*^*′*^) estimate by 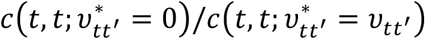 to adjust it for sample overlap.

### Real data application

We estimated shared and non-shared disease associated gene counts using gene-based test statistics (*T*_*k*_) for 32 complex traits and statements of their null and non-null distributions downloaded from a public database [20]. A full list of the repositories from which these GWAS data were accessed is available in the **Appendix**, where the phenotype label abbreviations and GWAS sample sizes are also present. All GWAS were performed in populations of predominantly or exclusively European ancestry.

Genetic correlation estimates using LDSC are presented for all 496 trait pairs in **Supplementary Figure S11**. Gene-based test statistics were calculated as the sum of SNP-level association chi-square statistics from GWAS for all SNPs within ±50 Kb of the gene start and end base pair positions defined using Ensembl [73], and which could be matched to the 1000 Genome Phase 3 European (1KGv3-EUR) LD reference panel [74]. We defined Bonferroni significant trait-genes as those with a P-value less than 0.05/*d*, where *d* is the number of independent gene-based association test statistics estimated using the method of [75] applied to each chromosome separately and summed across them. Let **Σ**_*c*_ be the *R*_*c*_ × *R*_*c*_ matrix of correlations between gene-based test statistics for a single trait calculated using the method described previously for the *c*th chromosome and where 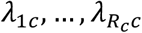 are its eigenvalues. We calculated *d*_*c*_ as

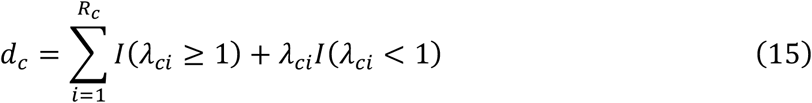

where *I*(*a*) is 1 if the argument *a* is true and 0 otherwise. Where any **Σ**_*c*_ matrix was not positive definite, we used the nearest positive definite matrix via the transform of [76]. The number of independent gene-based association tests performed across the genome was approximated as *d* = ∑_*c*_ *d*_*c*_ which we found to be 12,272 using the 1KGv3-EUR reference. LD scores and minor allele frequencies which parameterized the SNP-level and subsequent gene-level models were calculated using the 1KGv3-EUR reference panel. LD scores were calculated using SNPs in windows 1 centimorgan wide.

To count the numbers of independent Bonferroni- and false discovery rate (FDR)- significant genes in gene-based association testing as we show in **Figure 1a**, we first formed the set {*T*_*𝓁*_} such that *T*_*𝓁*_ < *T*_*s*_ ∀ *s* ∈ {*s*: *s* ≠ *𝓁*, Corr(*T*_*𝓁*_, *T*_*s*_) < √0.5}. The number of independent significant genes was then the size of the set {*T*_*𝓁*_} using either Bonferroni or FDR correction. To identify drug target candidates, we joined lists of drug-target interactions from ChEMBL [77], BindingDB [78], and GtoPdb [79] to their molecules indicated in DrugBank [80], of which there were 3,369. We also estimated the risk-directed genetic correlation between traits (*cf*. Equation 9), and all traits (see **Appendix**) were inferred to already be coded in the direction of risk in their respective GWAS except HDL, intelligence, and education.

## Appendix

Here we present the names of each trait we studied, its abbreviation displayed throughout the manuscript, the PubMed ID to the study from which the original GWAS data came, and the GWAS sample size using the convention of <**abbreviation**: Full name (PubMed ID; GWAS sample size)>. All GWAS cohorts were of strictly or predominantly European ancestry.

- **AD**: Alzheimer’s disease (35379992; 487,511)
- **ADHD**: Attention-deficit/hyperactivity disorder (36702997; 225,534)
- **AFIB**: Atrial fibrillation (36653681; 2,339,188)
- **agingR1**: R1 index of aging-associated brain atrophy (39147830; 49,482)
- **agingR2**: R2 index of aging-associated brain atrophy (39147830; 49,482)
- **agingR3**: R3 index of aging-associated brain atrophy (39147830; 49,482)
- **agingR4**: R4 index of aging-associated brain atrophy (39147830; 49,482)
- **agingR5**: R5 index of aging-associated brain atrophy (39147830; 49,482)
- **ALS**: Amyotrophic lateral sclerosis (34873335; 138,086)
- **BIP**: Bipolar I or II disorder (34002096; 413,466)
- **BMI**: Body mass index (30239722; 694,649)
- **CAD**: Coronary artery disease (36474045; 1,165,690)
- **CKD**: Chronic kidney disease (31152163; 625,219)
- **DBP**: Diastolic blood pressure (30224653; 757,601)
- **EDU**: Educational attainment (35361970; 3,037,499)
- **HDL**: High-density lipoprotein cholesterol (34887591; 1,320,016)
- **INT**: Fluid intelligence (36150907; 216,381)
- **LBD**: Lewy body dementia (33589841; 16,516)
- **LDL**: Low-density lipoprotein cholesterol (34887591; 1,320,016)
- **LUPUS**: Lupus (34278373; 324,698)
- **MDD**: Major depressive disorder (30718901; 807,553)
- **MS**: Multiple sclerosis (34737426; 456,348)
- **mvAGING**: Multivariate index of non-pathologic aging (37550455; 1,958,000)
- **PD**: Parkinson’s disease (38155330; 611,485)
- **PP**: Pulse pressure (30224653; 757,601)
- **RA**: Rheumatoid arthritis (34737426; 456,348)
- **SBP**: Systolic blood pressure (30224653; 757,601)
- **SCZ**: Schizophrenia (35396580; 320,404)
- **SLEEP**: Sleep duration (30846698; 446,118)
- **STROKE**: Ischemic stroke (36180795; 1,296,908)
- **TG**: Triglycerides (34887591; 1,320,016)
- **T2D**: Type 2 diabetes (37034649; 1,528,967)

## Software availability

We developed an R package to compute gene-level posterior risk probabilities and estimates of shared polygenicity between traits which requires only gene- or SNP-level summary statistics from GWAS and an LD reference panel (1000 Genomes Phase 3 is provided) at https://github.com/noahlorinczcomi/bpact. All R code used to perform the analyses presented in the main text is available at https://github.com/noahlorinczcomi/bpact_analysis.

## Data Availability

All data used in this study was downloaded from a public database of gene-based test statistics for complex traits: https://nlorinczcomi.shinyapps.io/gent/. Linkage disequilibrium reference panels were from the European population of 1000 Genomes Phase 3 study [74] available at https://www.internationalgenome.org/ or https://github.com/privefl/bigsnpr. We provide posterior risk probabilities for each of the 32 traits we studied and up to 17,166 genes and make the results available at https://github.com/noahlorinczcomi/bpact. We also provide estimated total, shared, and non-shared disease associated gene counts in the **Supplemental Data**.

## References

1. Visscher, P.M., et al., Discovery and implications of polygenicity of common diseases. Science, 2021. 373(6562): p. 1468–1473.

2. Jiang, X., et al., Shared heritability and functional enrichment across six solid cancers. Nature communications, 2019. 10(1): p. 431.

3. Ballard, J.L. and L.J. O’Connor, Shared components of heritability across genetically correlated traits. The American Journal of Human Genetics, 2022. 109(6): p. 989–1006.

4. Lounkine, E., et al., Large-scale prediction and testing of drug activity on side-effect targets. Nature, 2012. 486(7403): p. 361–367.

5. Bedi, O., et al., Pleiotropic effects of statins: new therapeutic targets in drug design. Naunyn-Schmiedeberg’s archives of pharmacology, 2016. 389: p. 695–712.

6. Nguyen, P.A., et al., Phenotypes associated with genes encoding drug targets are predictive of clinical trial side effects. Nature communications, 2019. 10(1): p. 1579.

7. Woodward, D.J., et al., Leveraging pleiotropy for the improved treatment of psychiatric disorders. Molecular Psychiatry, 2024: p. 1–17.

8. Mao, T., et al., Causal relationships between GLP1 receptor agonists, blood lipids, and heart failure: a drug-target mendelian randomization and mediation analysis. Diabetology & Metabolic Syndrome, 2024. 16(1): p. 208.

9. Horton, J.D., J.C. Cohen, and H.H. Hobbs, Molecular biology of PCSK9: its role in LDL metabolism. Trends in biochemical sciences, 2007. 32(2): p. 71–77.

10. Cohen, J.C.B. E.; Mosley Jr, T. H.; Hobbs, H. H., Sequence variations in PCSK9, low LDL, and protection against coronary heart disease. New England Journal of Medicine, 2006. 354(12): p. 1264–1272.

11. Leeuw, C.A., et al., MAGMA: generalized gene-set analysis of GWAS data. PLoS computational biology, 2015. 11(4): p. 1004219.

12. Sivakumaran, S., et al., Abundant pleiotropy in human complex diseases and traits. The American Journal of Human Genetics, 2011. 89(5): p. 607–618.

13. Bulik-Sullivan, B.K., et al., Schizophrenia Working Group of the Psychiatric Genomics Consortium. Nature genetics, 2015. 47(3): p. 291–295.

14. Wu, T., et al., Polygenic power calculator: Statistical power and polygenic prediction accuracy of genome-wide association studies of complex traits. Frontiers in Genetics, 2022. 13: p. 989639.

15. Schaid, D.J., et al., Statistical methods for testing genetic pleiotropy. Genetics, 2016. 204(2): p. 483–497.

16. Frei, O.H. D.; Smeland, O.B.; Shadrin, A.A.; Fan, C.C.; Maeland, S.; O’Connell, K.S.; Wang, Y.; Djurovic, S.; Thompson, W.K.; Andreassen, O.A.;, Bivariate causal mixture model quantifies polygenic overlap between complex traits beyond genetic correlation. Nature communications, 2019. 10(1).

17. Parker, N.C. W.; Hindley, G.F.; O’Connell, K.S.; Parekh, P.; Hagen, E.; Smeland, O.B.; Djurovic, S.; Shadrin, A.A.; Andreassen, O.A.; Frei, O., Sourcing Bivariate Genetic Overlap for Polygenic Prediction using MiXeR-Pred. medrXiv, 2024.

18. Akdeniz, B.C.F. O.; Shadrin, A.; Vetrov, D.; Kropotov, D.; Hovig, E.; Andreassen, O.A.; Dale, A.M., Finemap-MiXeR: A variational Bayesian approach for genetic finemapping. PLoS Genetics, 2024. 20(8).

19. Stefanski, L.A. and J.R. Cook, Simulation-extrapolation: the measurement error jackknife. Journal of the American Statistical Association, 1995. 90(432): p. 1247–1256.

20. Lorincz-Comi, N.S. W.; Chen, X.; Rivera Paz, I.; Hou, Y.; Zhou, Y.; Xu, J.; William, M.; Barnard, J.; Pieper, A.; Haines, J.; Chung, M.; Cheng, F., Combining xQTL and Genome-Wide Association Studies from Ethnically Diverse Populations Improves Druggable Gene Discovery. SRN, 2025.

21. Sachse, S.M., et al., Nuclear import of the DSCAM-cytoplasmic domain drives signaling capable of inhibiting synapse formation. The EMBO journal, 2019. 38(6): p. 99669.

22. Chen, X., et al., The study of Alzheimer’s disease risk diagnosis based on natural killer cell marker genes in the multi-omics data. Journal of Alzheimer’s Disease, 2024. 0(0).

23. Ballantyne, C.M., Low-density lipoproteins and risk for coronary artery disease. The American journal of cardiology, 1998. 82(8): p. 3–12.

24. Xie, J., et al., The unsupervised feature selection algorithms based on standard deviation and cosine similarity for genomic data analysis. Frontiers in Genetics, 2021. 12: p. 684100.

25. Frech, C. and N. Chen, Genome-wide comparative gene family classification. PLoS one, 2010. 5(10): p. 13409.

26. Rosoff, D.B., et al., Multivariate genome-wide analysis of aging-related traits identifies novel loci and new drug targets for healthy aging. Nature aging, 2023. 3(8): p. 1020–1035.

27. Qiang, Y.X., et al., Associations of blood cell indices and anemia with risk of incident dementia: A prospective cohort study of 313,448 participants. Alzheimer’s & Dementia, 2023. 19(9): p. 3965–3976.

28. Arbogast, T., et al., Kctd13-deficient mice display short-term memory impairment and sex-dependent genetic interactions. Human molecular genetics, 2019. 28(9): p. 1474–1486.

29. Jadiya, P., et al., Impaired mitochondrial calcium efflux contributes to disease progression in models of Alzheimer’s disease. Nature communications, 2019. 10(1): p. 3885.

30. Jadiya, P., et al., Genetic Rescue of Mitochondrial Calcium Efflux in Alzheimer’s Disease Preserves Mitochondrial Function and Protects against Neuronal Cell Death. Biophysical Journal, 2017. 112(3): p. 445.

31. Lonsdale, J., et al., The genotype-tissue expression (GTEx) project. Nature genetics, 2013. 45(6): p. 580–585.

32. Lorincz-Comi, N., et al., MRBEE: A bias-corrected multivariable Mendelian randomization method. Human Genetics and Genomics Advances, 2024. 5(3).

33. Mattiola, I., A. Mantovani, and M. Locati, The tetraspan MS4A family in homeostasis, immunity, and disease. Trends in Immunology, 2021. 42(9): p. 764–781.

34. Luo, X., et al., MS4A superfamily molecules in tumors, Alzheimer’s and autoimmune diseases. Frontiers in Immunology, 2024. 15: p. 1481494.

35. Ma, J., J.T. Yu, and L. Tan, MS4A cluster in Alzheimer’s disease. Molecular neurobiology, 2015. 51: p. 1240–1248.

36. Deming, Y., et al., The MS4A gene cluster is a key modulator of soluble TREM2 and Alzheimer’s disease risk. Science translational medicine, 2019. 11(505): p. 2291.

37. Falgarone, G. and G. Chiocchia, Clusterin: A multifacet protein at the crossroad of inflammation and autoimmunity. Advances in cancer research, 2009. 104: p. 139–170.

38. Sardi, F., et al., Alzheimer’s disease, autoimmunity and inflammation. The good, the bad and the ugly. Autoimmunity reviews, 2011. 11(2): p. 149–153.

39. Bergink, V., S.M. Gibney, and H.A. Drexhage, Autoimmunity, inflammation, and psychosis: a search for peripheral markers. Biological psychiatry, 2014. 75(4): p. 324–331.

40. Jong, M.C., M.H. Hofker, and L.M. Havekes, Role of ApoCs in lipoprotein metabolism: functional differences between ApoC1, ApoC2, and ApoC3. Arteriosclerosis, thrombosis, and vascular biology, 1999. 19(3): p. 472–484.

41. Gao, M., et al., ApoC2 deficiency elicits severe hypertriglyceridemia and spontaneous atherosclerosis: A rodent model rescued from neonatal death. Metabolism, 2020. 109: p. 154296.

42. Knox, C., et al., DrugBank 6.0: the DrugBank knowledgebase for 2024. Nucleic acids research, 2024. 52(D1): p. 1265–1275.

43. Tomar, M., et al., A clinical and computational study on anti-obesity effects of hydroxycitric acid. RSC advances, 2019. 9(32): p. 18578–18588.

44. Yadikar, N., et al., Exploring the mechanism of citric acid for treating glucose metabolism disorder induced by hyperlipidemia. Journal of Food Biochemistry, 2022. 46(12): p. 14404.

45. Amin, K.A., H.H. Kamel, and M.A. Abd Eltawab, The relation of high fat diet, metabolic disturbances and brain oxidative dysfunction: modulation by hydroxy citric acid. Lipids in health and disease, 2011. 10: p. 1–11.

46. Abdel-Salam, O.M.E., et al., Citric acid effects on brain and liver oxidative stress in lipopolysaccharide-treated mice. Journal of medicinal food, 2014. 17(5).

47. Suner, S.S., et al., Versatile fluorescent carbon dots from citric acid and cysteine with antimicrobial, anti-biofilm, antioxidant, and AChE enzyme inhibition capabilities. Journal of fluorescence, 2021. 31(6): p. 1705–1717.

48. Marucci, G., et al., Efficacy of acetylcholinesterase inhibitors in Alzheimer’s disease. Neuropharmacology, 2021. 190: p. 108352.

49. Cerbai, F., et al., N1phenethyl-norcymserine, a selective butyrylcholinesterase inhibitor, increases acetylcholine release in rat cerebral cortex: a comparison with donepezil and rivastigmine. European journal of pharmacology, 2007. 572(2-3): p. 142–150.

50. Furukawa-Hibi, Y., et al., Butyrylcholinesterase inhibitors ameliorate cognitive dysfunction induced by amyloid-β peptide in mice. Behavioural brain research, 2011. 225(1): p. 222–229.

51. Markham, A., Fostamatinib: first global approval. Drugs, 2018. 78: p. 959–963.

52. Yergolkar, A.V., et al., PND24 target identification and drug repurposing for Parkinson’s disease: A novel integrative computational approach. Value in Health Regional Issues, 2020. 22: p. 79.

53. Duan, Q.Q., et al., TBK1, a prioritized drug repurposing target for amyotrophic lateral sclerosis: evidence from druggable genome Mendelian randomization and pharmacological verification in vitro. BMC medicine, 2024. 22(1): p. 96.

54. Eshak, D. and M. Arumugam, Unveiling therapeutic biomarkers and druggable targets in ALS: An integrative microarray analysis, molecular docking, and structural dynamic studies. Computational Biology and Chemistry, 2024. 113: p. 108211.

55. Birkle, T.J. and G.C. Brown, Syk inhibitors protect against microglia-mediated neuronal loss in culture. Frontiers in Aging Neuroscience, 2023. 15: p. 1120952.

56. Zhou, C., et al., Activation of spleen tyrosine kinase (SYK) contributes to neuronal pyroptosis and cognitive impairment in diabetic mice via the NLRP3/Caspase-1/GSDMD signaling pathway. Experimental Gerontology, 2024. 198: p. 112626.

57. Miyazaki, M., et al., Targeted disruption of GAK stagnates autophagic flux by disturbing lysosomal dynamics. International Journal of Molecular Medicine, 2021. 48(4): p. 1–18.

58. Pan, J., et al., A kinome-wide siRNA screen identifies multiple roles for protein kinases in hypoxic stress adaptation, including roles for IRAK4 and GAK in protection against apoptosis in VHL−/− renal carcinoma cells, despite activation of the NF-κB pathway. Journal of biomolecular screening, 2013. 18(7): p. 782–796.

59. Liu, T., et al., NF-κB signaling in inflammation. Signal transduction and targeted therapy, 2017. 2(1): p. 1–9.

60. Shi, J.H., X. Xie, and S.C. Sun, TBK1 as a regulator of autoimmunity and antitumor immunity. Cellular & molecular immunology, 2018. 15(8): p. 743–745.

61. Balka, K.R., et al., TBK1 and IKKε act redundantly to mediate STING-induced NF-κB responses in myeloid cells. Cell reports, 2020. 31(1).

62. Qian, Y., et al., The sodium channel subunit SCNN1B suppresses colorectal cancer via suppression of active c-Raf and MAPK signaling cascade. Oncogene, 2023. 42(8): p. 601–612.

63. Sahana, T.G. and K. Zhang, Mitogen-activated protein kinase pathway in amyotrophic lateral sclerosis. Biomedicines, 2021. 9(8): p. 969.

64. Yadav, R.K., E. Minz, and S. Mehan, Understanding abnormal c-JNK/p38MAPK signaling in amyotrophic lateral sclerosis: potential drug targets and influences on neurological disorders. CNS & Neurological Disorders-Drug Targets (Formerly Current Drug Targets-CNS & Neurological Disorders, 2021. 20(5): p. 417–429.

65. Lemmon, M.A., Pleckstrin homology (PH) domains and phosphoinositides, in Biochemical Society Symposia. 2007, Portland Press Ltd. p. 81–93.

66. Bellenguez, C.K. F.; Jansen, I.E.; Kleineidam, L.; Moreno-Grau, S.; Amin, N.; Naj, A.C.; Campos-Martin, R.; Grenier-Boley, B.; Andrade, V.; Holmans, P.A., New insights into the genetic etiology of Alzheimer’s disease and related dementias. Nature genetics, 2022. 54(4): p. 412–436.

67. Loos, R.J. and G.S. Yeo, The genetics of obesity: from discovery to biology. Nature Reviews Genetics, 2022. 23(2): p. 120–133.

68. Wu, T. and P.C. Sham, On the transformation of genetic effect size from logit to liability scale. Behavior Genetics, 2021. 51(3): p. 215–222.

69. Zhu, X. and M. Stephens, Bayesian large-scale multiple regression with summary statistics from genome-wide association studies. The annals of applied statistics, 2017. 11(3): p. 1561.

70. Privé, F., Optimal linkage disequilibrium splitting. Bioinformatics, 2022. 38(1): p. 255–256.

71. Rubin, D.B., Multiple imputation after 18+ years. Journal of the American statistical Association, 1996. 91(434): p. 473–489.

72. LeBlanc, M., et al., Schizophrenia and Bipolar Disorder Working Groups of the Psychiatric Genomics Consortium. BMC genomics, 2018. 19: p. 1–15.

73. Harrison, P.W., et al., Ensembl 2024. Nucleic acids research, 2024. 52(D1): p. 891–899.

74. Siva, N., 1000 Genomes project. Nature biotechnology, 2008. 26(3): p. 256–257.

75. Jiang, L., et al., Powerful and robust inference of complex phenotypes’ causal genes with dependent expression quantitative loci by a median-based Mendelian randomization. The American Journal of Human Genetics, 2022. 109(5): p. 838–856.

76. Choi, Y.G., et al., Fixed support positive-definite modification of covariance matrix estimators via linear shrinkage. Journal of Multivariate Analysis, 2019. 171: p. 234–249.

77. Gaulton, A., et al., ChEMBL: a large-scale bioactivity database for drug discovery. Nucleic acids research, 2012. 40(D1): p. 1100–1107.

78. Liu, T., et al., BindingDB in 2024: a FAIR knowledgebase of protein-small molecule binding data. Nucleic Acids Research, 2024. gkae1075.

79. Harding, S.D., et al., The IUPHAR/BPS guide to PHARMACOLOGY in 2022: curating pharmacology for COVID-19, malaria and antibacterials. Nucleic Acids Research, 2022. 50(D1): p. 1282–1294.

80. Wishart, D.S., et al., DrugBank 5.0: a major update to the DrugBank database for 2018. Nucleic acids research, 2018. 46(D1): p. 1074–1082.

