## Supplement for "Bayesian inference of genetic pleiotropy identifies drug targets and repurposable medicines for human complex diseases"

### Moment matching distributions of GenT test statistics under SNP model

We show in this section how the null and null distributions of the gene-based test statistics using GenT (Lorincz-Comi et al., 2024a) are parameterized under the random effect SNP-level model we introduced in the main text. Let $T_k$ represent the gene-based test statistic for the $k$th gene, $\mathbf{z}_k = \left(\sqrt{N}\sigma_j\hat{\beta}_j^k\right)$ be the vector of Z-statistics for $m_k$ SNPs used to test the $k$th gene, $\mathbf{L}_k = \mathrm{diag}\left(\ell_j\right)$ be the diagonal matrix of LD scores for these SNPs, $\mathbf{D}_k = \mathrm{diag}\left(\sigma_j^2\right)$ be the diagonal matrix of $\mathrm{E}(G_{ik}^2)$ values, and $\mathbf{H}_k \coloneqq \mathbf{D}_k^{1/2}\mathbf{L}_k\mathbf{D}_k^{1/2}$ such that $\mathrm{Cov}(\mathbf{z}_k|H_{1k}) = N\tau\mathbf{H}_k + \mathbf{R}_k$ and $\mathrm{Cov}(T_k|H_{0k}) = \mathbf{R}_k$, where $\mathbf{R}_k$ is the LD matrix, or $\mathrm{Corr}(\mathbf{z}_k)$, and $H_{0k}\colon \mathrm{E}(T_k) = m_k$ and $H_{1k}\colon \mathrm{E}(T_k) > m_k$. It follows that

$$\mathrm{E}(T_k|H_{0k}) = m_k$$
$$\mathrm{Var}(T_k|H_{0k}) = 2\mathrm{tr}(\mathbf{R}_k\mathbf{R}_k)$$

and

$$\begin{aligned}
\mathrm{E}(T_k|H_{1k}) &= \mathrm{tr}[\mathrm{Cov}(\mathbf{z}_k|H_{1k})] \\
&= m_k + N\tau\sum_{j=1}^{m_k}\sigma_j^2\ell_j\,, \\
\mathrm{Var}(T_k|H_{1k}) &= 2\mathrm{tr}[(\tau\mathbf{H}_kN + \mathbf{R}_k)(\tau\mathbf{H}_kN + \mathbf{R}_k)] \\
&= 2\mathrm{tr}(\mathbf{R}_k\mathbf{R}_k) + 2(N\tau)^2\sum_{j=1}^{m_k}\sigma_j^4\ell_j^2 + 4N\tau\sum_{j=1}^{m_k}\sigma_j^2\ell_j
\end{aligned}$$

since $\mathrm{tr}(\mathbf{H}_k\mathbf{R}_k) = \mathrm{tr}(\mathbf{H}_k)$ because $\mathbf{H}_k$ is a diagonal matrix and $\mathbf{R}_k$ is a correlation matrix. These quantities are used to parameterize the non-null distribution of $T_k$ using the method of moments with the Gamma distribution, namely by $\xi_{0k} = \mathrm{E}(T_k|H_{0k})\mathrm{Var}(T_k|H_{0k})^{-1}$, $\alpha_{0k} = \mathrm{E}(T_k|H_{0k})\xi_{0k}$, $\xi_{1k} = \mathrm{E}(T_k|H_{1k})\mathrm{Var}(T_k|H_{1k})^{-1}$ and $\alpha_{1k} = \mathrm{E}(T_k|H_{1k})\xi_{1k}$ where $T_k|H_{0k} \overset{\text{approx.}}{\sim} \Gamma(\alpha_{0k}, \xi_{0k})$ and $T_k|H_{1k} \overset{\text{approx.}}{\sim} \Gamma(\alpha_{1k}, \xi_{1k})$ (Lorincz-Comi et al., 2024).

### Convergence of gene-specific posterior causal probabilities

We attempt to show that the gene-specific posterior causal probability for the $k$th gene and $t$th trait based only on the posterior likelihood, denoted here as

$$p = \frac{\mathrm{f}_1(T)(1-\delta)}{\mathrm{f}_1(T)(1-\delta) + \mathrm{f}_0(T)\delta}\,,$$

without subscripts for notational convenience, where $T$ is the gene-based test statistic using $m$ SNPs, $\mathrm{f}_0$ and $\mathrm{f}_1$ its distribution under $H_0\colon \mathrm{E}(T) = m$ and $H_1\colon \mathrm{E}(T) > m$, and prior causal probability $(1-\delta)$, converges to 1 when the gene is causal, i.e., when the average scaled SNP heritability $\tau > 0$. Since $0 < \delta < 1$ is a constant that does affect the convergence of $p$, only its rate, we ignore it. Since we intend to demonstrate this relationship for a likelihood inference regarding the $k$th gene, we will use a fixed effect model for SNP effects instead of the random effects model introduced in the main text for genome-wide gene sets. Stating that $p \to 1$ as $n \to \infty$ is equivalent to demonstrating that

$$\lim_{n\to\infty,\tau>0} \log\left[\left(\frac{\mathrm{f}_1(T_n)}{\mathrm{f}_0(T_n)}\right)\right] = 1,$$

where $n$ is the GWAS sample size, such that $T_n \coloneqq \mathbf{z}_n^\top \mathbf{z}_n$ when $\mathbf{z}_n = \sqrt{n}\widehat{\boldsymbol{\beta}}$, $\widehat{\boldsymbol{\beta}} = \left(\hat{\beta}_j\right)_{j=1}^{m}$ and $\mathrm{Corr}\left(\widehat{\boldsymbol{\beta}}\right) \coloneqq \mathbf{R}$. As mentioned in the main text, we used the GenT test statistics (Lorincz-Comi et al., 2024a) which follow $\Gamma(\alpha_0, \xi_0)$ under $H_0$ and $\Gamma(\alpha_{1n}, \xi_{1n})$ under $H_1$, where

$$\alpha_0 = \frac{m^2}{2\mathrm{tr}(\mathbf{RR})}$$
$$\xi_0 = \frac{m}{2\mathrm{tr}(\mathbf{RR})}$$
$$\alpha_{1n} = \frac{(m + n\boldsymbol{\beta}^\top\boldsymbol{\beta})^2}{2\mathrm{tr}(\mathbf{RR}) + 4n\boldsymbol{\beta}^\top\mathbf{R}\boldsymbol{\beta}}$$
$$\xi_{1n} = \frac{m + n\boldsymbol{\beta}^\top\boldsymbol{\beta}}{2\mathrm{tr}(\mathbf{RR}) + 4n\boldsymbol{\beta}^\top\mathbf{R}\boldsymbol{\beta}},$$

and we know have conditioned on $\boldsymbol{\beta}$, i.e. used a fixed instead of random effect model for $\boldsymbol{\beta}$, since we intend to make an inference about a specific gene. We can directly express $\log[\mathrm{f}_1(T_n)/\mathrm{f}_0(T_n)]$ as

$$\log\left[\left(\frac{\mathrm{f}_1(T_n)}{\mathrm{f}_0(T_n)}\right)\right] = \alpha_{1n}\log(\xi_{1n}) + (\alpha_{1n} - 1)\log(T_n) - \xi_{1n}T_n - \log[(\alpha_{1n} - 1)!] - \alpha_0\log(\xi_0) - (\alpha_0 - 1)\log(T_n) + \beta_0 T_n + \log[(\alpha_0 - 1)!]$$

Under $H_1$, $T_n$ is of the same asymptotic order as $n$ (i.e., $T_n \sim n$). As a result, the quantity above is of the same order as

$$n\log(n) - nd_1\log(n),$$

which is equal to

$$= n\log(n)\,(1 - d_1) \to \infty \text{ as } n \to \infty$$

under the conditions that $\alpha_0, \xi_0$, and $\xi_1$ are each converging to constants asymptotically as $n \to \infty$ and where

$$\alpha_{1n} \sim d_1 n,$$

$$d_1 = \frac{(\boldsymbol{\beta}^\top \boldsymbol{\beta})^2}{4\boldsymbol{\beta}^\top \mathbf{R}\boldsymbol{\beta}}.$$

The result also follows from the fact that $d_{1n} < 1$ since $(\boldsymbol{\beta}^\top \boldsymbol{\beta})^2 < \boldsymbol{\beta}^\top \boldsymbol{\beta} < \boldsymbol{\beta}^\top \mathbf{R}\boldsymbol{\beta} < 4\boldsymbol{\beta}^\top \mathbf{R}\boldsymbol{\beta}$**.** That

$$\lim_{n\to\infty,\tau>0} \log\left[\left(\frac{\mathrm{f}_1(T_n)}{\mathrm{f}_0(T_n)}\right)\right] = 1$$

suggests $p \to 1$ as $n \to \infty$.

### Bias analysis of estimated number of disease associated genes

In this section, we show how we arrived at the quantity representing bias in our estimated of the number of disease associated genes for a single trait in the main text. It was shown in the main text that our estimate $c(t)$ of the number of causal genes for a trait is the sum of posterior probabilities across $M$ tested genes that each binary indicator $I_k$ takes value 1. Let $I_k^0$ denote represent the true binary indicator of causality for the $k$th gene, of which $I_k$ is an estimate. Using the notation from the main text, let $\ddot{M} = \sum_k I_k^0 = M(1-\delta)$ represent the true number of causal genes. Our estimator $c(t)$ has the following relationship with $\ddot{M}$:

$$c(t) \coloneqq \sum_k \mathrm{P}(I_k = 1|T_k)$$

where $T_k$ is the gene-based test statistic for the $k$th gene and the posterior probability is taken after integration of the parameters $(\delta, \tau)$ defining the polygenic model of the trait. In finite GWAS samples, it follows that

$$\begin{aligned}
\mathrm{E}[c(t)] &= \mathrm{E}\left[\sum_{k=1}^{M} \mathrm{P}(I_k = 1|T_k)\right] \\
&= \mathrm{E}\left[\sum_{k=1}^{M} \mathrm{E}(I_k|T_k)\right] \\
&= \sum_{k=1}^{M} \mathrm{E}(I_k) \\
&= \sum_{s=0}^{1}\sum_{k=1}^{M} \mathrm{E}(I_k \middle| I_k^0 = s)\mathrm{P}(I_k^0 = s) \\
&= \sum_{s=0}^{1}\sum_{k=1}^{M} \mathrm{P}(I_k = 1 \middle| I_k^0 = s)\mathrm{P}(I_k^0 = s).
\end{aligned}$$

We can expand the final term as

$$(1-\delta)\sum_{k=1}^{M}\underbrace{\mathrm{P}(I_k=1|I_k^0=1)}_{\text{sensitivity}}+\delta\sum_{k=1}^{M}\underbrace{\mathrm{P}(I_k=1|I_i^0=0)}_{1-\text{specificity}}$$

where the sensitivity and (1-specificity) values are from using the indicator $I_k$ to estimate causality. In infinite GWAS samples, the result from the previous section suggests that the sensitivity will be 1.

We performed a simulation to evaluate the precision of $c(t)$ in estimating $\ddot{M}$ as $\ddot{M}$, heritability, and GWAS sample size changed by generating data under the SNP-level model introduced in the main text and applying $c(t)$ with known and fixed $(\delta,\tau)$ values, denoted here as $c_{\delta,\tau}(t)$. The data-generating process follows that described when the SIMEX adjustment for GWAS sample overlap was introduced in the main text, and heritability and GWAS sample sizes were chosen to align with the values we saw in our real data analyses. These results displayed in **Figure S1** suggest that $c_{\delta,\tau}(t)$ is very unlikely to be inflated relative to the true number of causal genes, and that underestimation of $\ddot{M}$ by $c_{\delta,\tau}(t)$ is only likely when GWAS are underpowered, i.e., when the ratio of GWAS sample size is small, SNP heritability is small, and the number of true causal genes is large. Otherwise, $c_{\delta,\tau}(t)$ can be highly precise in many scenarios.

**Figure S1: Estimation performance of $c(t)$**

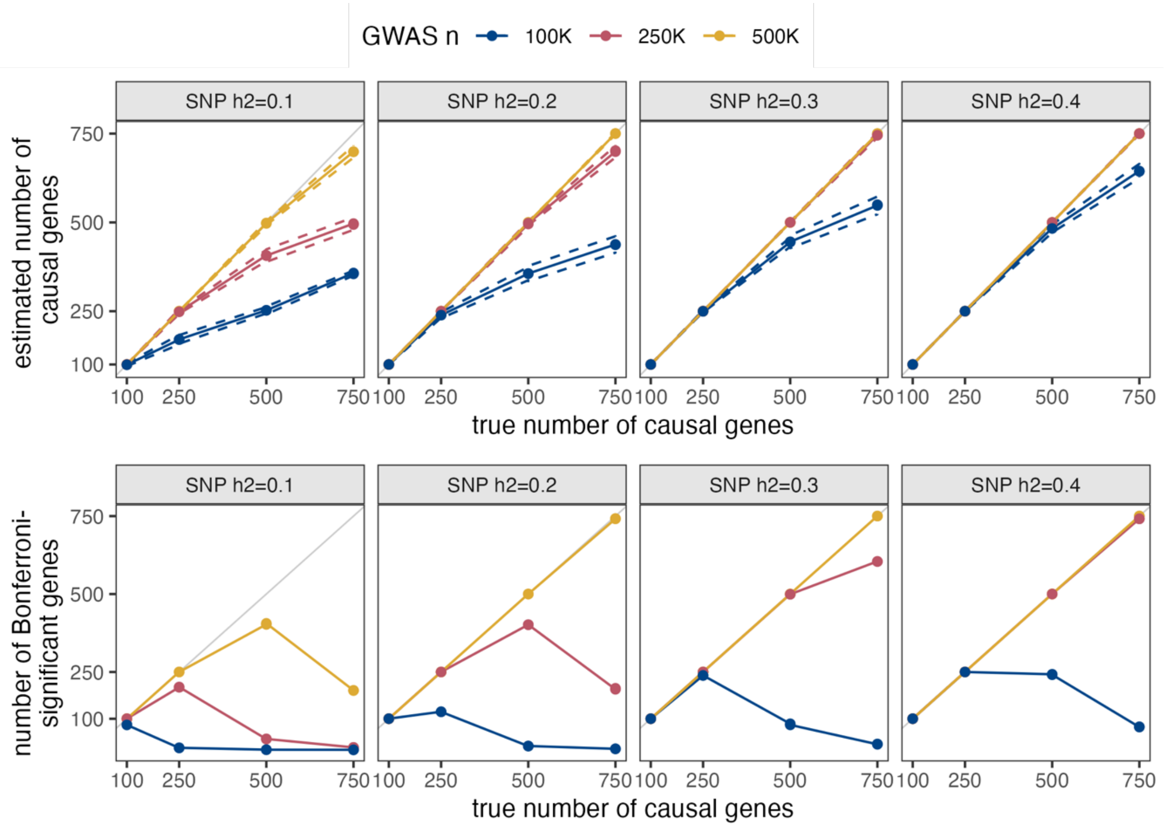

(top) Each point is the average of estimated causal gene counts across 20 simulations and dashed lines are plus/minus 2 times their standard deviation. The diagonal gray line represents y=x. (bottom) Each point is the number of Bonferroni-significant (0.05/12,727; the threshold used in practice during genome-wide hypothesis testing) gene-based association test statistics. The complete R code that we used to produce these results is available at https://github.com/noahlorinczcomi/bpact.

### Estimating posteriors distributions of model parameters $\delta$ and $\tau$

#### Metropolis-Hastings algorithm

In practice, we sample from the posterior distributions $p_{\delta_0}(\delta|T_k)$ and $p_{\tau_0}(\tau|T_k)$ using the Metropolis-Hastings (MH) algorithm (Chib & Greenberg, 1995) and approximate the posterior $p_{\delta_0}(\delta|T_k)$ by moment-matching it to a Beta distribution and approximate $p_{\tau_0}(\tau|T_k)$ by searching a grid of Trapezoidal parameters which most closely matches the empirical cumulative density function of posterior replicates from MH. The priors of these distributions respectively are Beta(100,1+1E-4) and Trapezoidal(2E-10,2E-9,2E-7,2E-5) and

$$
\begin{aligned}
f(T_k^t|\delta^t,\tau^t) &\sim f(T_k^t|\delta^t,\tau^t,I_k^t=1)P(I_k^t=1|\delta^t,\tau^t)+f(T_k^t|\delta^t,\tau^t,I_k^t=0)P(I_k^t=0|\delta^t,\tau^t)\\
&\sim \delta^t\Gamma(\alpha_{0k}^t,\xi_{0k}^t)+(1-\delta^t)\Gamma(\alpha_{1k}^t,\xi_{1k}^t)
\end{aligned}
$$

using the notation from above for $(\alpha_{0k}^t, \xi_{ok}^t, \alpha_{1k}^t, \xi_{1k}^t)$. We perform the double integration introduced over the prior distributions of $\delta^t$ and $\tau^t$ that was introduced in the main text using a two-dimensional quadrature in the `pracma` R package (Borchers & Borchers, 2019). The full R code we use to perform this sampling is available at https://github.com/noahlorinczcomi/polgene.

### Correlation of test statistics within genes between GWAS cohorts

We show the correlation between gene-based test statistics from the same sets of gene-specific SNPs and two traits $t$ and $t'$, denoted as $T_k^t$ and $T_k^{t'}$ from GWAS studies of sizes $N_t$ and $N_{t'}$ subjects, of which $N_{tt'}$ are shared. We can state the multivariate normal distribution of $\mathbf{z}_k^t$ and $\mathbf{z}_k^{t'}$ under their respective null hypotheses as

$$\tilde{\mathbf{z}}(k; t, t') \coloneqq \begin{pmatrix} \mathbf{z}_k^t \\ \mathbf{z}_k^{t'} \end{pmatrix} \mid H_{0k}^t, H_{0k}^{t'} \sim N(\mathbf{0}, \boldsymbol{\Upsilon}_{tt'} \otimes \mathbf{R}_k)$$

where $\boldsymbol{\Upsilon}_{\mathrm{tt'}} = (\upsilon_{tt'})$ and $\upsilon_{tt'} \approx N_{tt'}(N_t N_{t'})^{-1/2}\mathrm{Corr}(t, t')$ is a correlation matrix. For the more general case of non-quantitative phenotypes $t$ and $t'$, the analogous factor $\upsilon_{tt'}$ is still proportional to the fraction of shared GWAS subjects between cohorts. It follows that $T_k^t + T_k^{t'} = \|\tilde{\mathbf{z}}(k; t, t')\|_2^2$ and that

$$\begin{aligned} \mathrm{Var}\left[\tilde{\mathbf{z}}(k; t, t')^\top \tilde{\mathbf{z}}(k; t, t') | H_{0k}^t, H_{0k}^{t'}\right] &= 2\mathrm{tr}[(\boldsymbol{\Upsilon}_{tt'} \otimes \mathbf{R}_k)(\boldsymbol{\Upsilon}_{tt'} \otimes \mathbf{R}_k)] \\ &= 2\mathrm{tr}(\boldsymbol{\Upsilon}_{tt'}\boldsymbol{\Upsilon}_{tt'})\mathrm{tr}(\mathbf{R}_k\mathbf{R}_k). \end{aligned}$$

Since $\mathrm{Var}\left(T_k^t + T_k^{t'} | H_{0k}^t, H_{0k}^{t'}\right) \coloneqq \mathrm{Var}(T_k^t) + \mathrm{Var}\left(T_k^{t'}\right) + 2\mathrm{Cov}\left(T_k^t, T_k^{t'}\right)$ and $\mathrm{Var}(T_k^t | H_{0k}^t) = \mathrm{Var}\left(T_k^{t'} \middle| H_{0k}^{t'}\right) = 2\mathrm{tr}(\mathbf{R}_k\mathbf{R}_k)$, it follows that

$$\mathrm{Corr}\left(T_k^t, T_k^{t'}\right) = \frac{1}{2}\mathrm{tr}(\boldsymbol{\Upsilon}_{tt'}\boldsymbol{\Upsilon}_{tt'}) - 1$$

which represents the correlation between gene-based test statistics from the same gene but calculated on two different traits from potentially different GWAS cohorts. This value is estimable from non-significant SNPs in each GWAS without having to calculate gene-based test statistics (Lorincz-Comi et al., 2024b). We can observe from this expression that the correlation between these statistics is strictly positive and depends entirely on the proportion of overlapping GWAS subjects and the phenotypic correlation between the traits.

### Relationship between number of significant and disease associated genes

We stated in the main text that the number of genome-wide significant genes in gene-based association testing for a single trait, scaled by the square root of the GWAS sample size, is approximately linearly proportional to the number of disease associated genes for the trait, and that linear correlation between the scaled number of significant genes and the estimated number of disease associated genes implies our estimated number of associated genes is at least linearly proportional to the number of truly associated genes. We will show in this section that the number of independent Bonferroni-significant genes scaled by the square root of the GWAS sample size $N$, denoted $\widehat{M}$, is proportional to the true number of disease associated genes $\ddot{M}$. We consider the gene-based test statistics from each of the $k$th genes for a single trait tested with $m_k$ SNPs that are used to test the null hypotheses $H_{0k}: \tau_k = 0$ vs $H_{1k}: \tau_k > 0$ across $M$ independent genes genome-wide at Bonferroni-corrected level $\alpha$, where now we define the gene-specific heritability explained $\tau_k$. Let $\gamma_k \coloneqq \mathrm{P}\left(T_k > Q_{k,\alpha} \middle| H_{1k}\right)$ where $Q_{k,\alpha}$ is such that $\mathrm{P}\left(T_k > Q_{k,a} \middle| H_{0k}\right) = \alpha$. We only consider genes in the set $\mathcal{T} = \{T_k\}$ formed such that $F_k(T_k) > F_s(T_s)\ \forall\ s \in \{s: s \neq k, \mathrm{Cov}(T_k, T_s) > \zeta\}$ where $\zeta$ is a threshold defining approximate non-independence set to 0.1 in practice, and $F_t$ is the cumulative density function of $t$. In the set $\mathcal{T}$, we define the indicator $I_k$ for each of its $M$ elements which takes the value 1 if $T_k > F_k^{-1}(1-\alpha)$ at significance level $\alpha$, and 0 otherwise. In other words, $I_k$ is a binary random variable indicating significance of a lead gene in gene-based association testing, and by definition

$$\widehat{M} = \sum_{k=1}^{M} I_k.$$

We can find $\mathrm{E}\left(\widehat{M}\right)$ as:

$$\begin{aligned}
\mathrm{E}\left(\widehat{M}\right) &= \sum_{k=1}^{M} \mathrm{E}(I_k) \\
&= \sum_{k=1}^{M} \mathrm{E}(I_k | H_{0k}) P(H_{0k}) + \mathrm{E}(I_k | H_{1k}) P(H_{1k}) \\
&= \sum_{k=1}^{M} \mathrm{P}(I_k = 1 | H_{0k}) \left(1 - \frac{\ddot{M}}{M}\right) + \mathrm{P}(I_k = 1 | H_{1k}) \frac{\ddot{M}}{M} \\
&= \left(1 - \frac{\ddot{M}}{M}\right) \sum_{k=1}^{M} \gamma_k + \ddot{M}\alpha \\
&= \ddot{M}\left(\frac{1}{M}\sum_{k=1}^{M} \gamma_k - \alpha\right) + M\alpha,
\end{aligned}$$

where $\gamma_k \coloneqq \mathrm{P}(I_k = 1 | H_{0k})$ and we replaced $\mathrm{P}(H_{0k})$ and $\mathrm{P}(H_{1k})$ respectively by their marginal probabilities $\delta \coloneqq 1 - \ddot{M}M^{-1}$ and $1 - \delta \coloneqq \ddot{M}M^{-1}$ from the main text. For simplicity, we can assume the power of each gene-based null hypothesis test is the same value, $\gamma$, and restate the above quantity as

$$\ddot{M}(\gamma - \alpha) + M\alpha.$$

Generally, $\alpha$ is a Bonferroni threshold defined such that $\alpha = \tilde{\alpha}/M$, where $\tilde{\alpha}$ is the nominal significance threshold. In this case, the above quantity further reduces to

$$\ddot{M}(\gamma - \alpha) + \tilde{\alpha}.$$

Under the further assumption that $g(\sqrt{N}) \coloneqq \gamma$ is approximately piecewise linear over fixed intervals $\sqrt{N}$ values (see **Figure S2**), we can state that $g(\sqrt{N}) \approx c\sqrt{N} + b$ in any of its branches, where $c$ and $b$ are generic constants that are branch-specific – but which are not denoted as such for notational simplicity. For example, in **Figure S2**, $b$ in the first branch is equal to $\alpha$ and in the last branch is equal to $1 - \alpha/2$. Using this approximation, it follows that

$$\ddot{M}(\gamma - \alpha) + \tilde{\alpha} \approx \ddot{M}\left(c\sqrt{N} + b - \alpha\right) + \tilde{\alpha}$$

which implies that

$$\frac{E(\hat{M})}{\sqrt{N}} \approx \ddot{M}\left(c + \frac{b - \alpha}{\sqrt{N}}\right) + \frac{\tilde{\alpha}}{\sqrt{N}} \propto \ddot{M},$$

which states that the expected number of Bonferroni significant and independent genes in gene-based association testing is approximately proportional to the true number of disease associated genes.

**Figure S2: Piecewise linear approximation of gene-based test power**

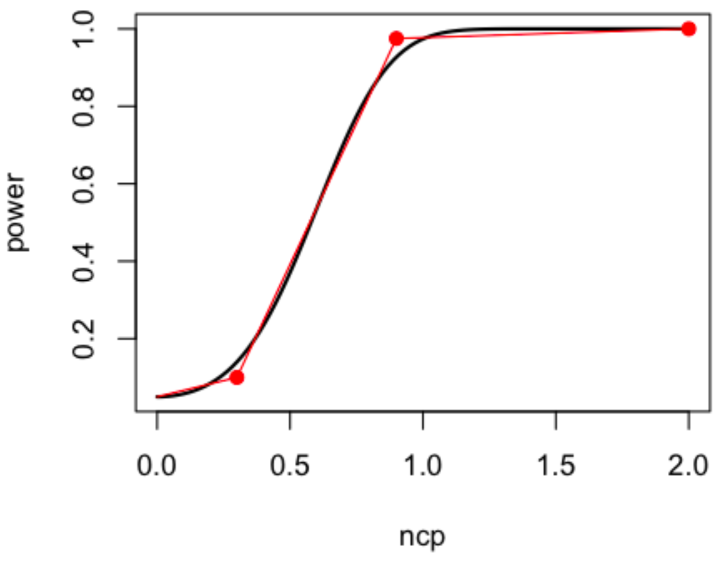

This result is from an analysis of power for a single gene tested with 100 SNPs that had a first-order autoregressive LD structure with correlation parameter 0.5. We tested non-centrality parameters (ncps) ranging from 0 to 2 and use the Gamma parameterization described in the **Methods** section of the main text and the first section of this **Supplement**. The ncps are proportional to the product of heritability and $\sqrt{N}$ in practice, and here we just consider their joint contribution for simplicity. For this example, displayed is the power is to reject the null hypothesis of the gene-based association test at the nominal significance level 0.05. The true power is denoted by the smooth black line and the piecewise linear function approximating it is denoted by red lines. Red points indicate the changepoints of the piecewise function. The full R code to reproduce these results is available at https://github.com/noahlorinczcomi/bpact.

### AD cosine/Jaccard index values and genetic correlations with 31 traits

We show scatterplots of cosine and Jaccard index values with the absolute values of estimated genetic correlation (LDSC; Bulik-Sullivan et al., 2015) between pairs of all 31 traits with AD in **Figure S3**. These results suggest no evidence of a linear correlation between these two index values and absolute genetic correlation across the 31 traits, but that many traits such as MDD, sleep duration, agingR3, BIP, LDL, TG, and LUPUS are estimated to share a nonzero proportion of disease associated genes but have very little evidence of a nonzero genetic correlation between them. This may be an artefact of the estimated genetic correlation which intends to only capture genome-wide correlation that is not sensitive to local shared genetic architectures. For example, the estimated genetic correlation may be approximately zero if an equal number of loci with strong negative and positive local genetic correlations is present for a pair of traits.

**Figure S3: Association between index values and genetic correlations with AD**

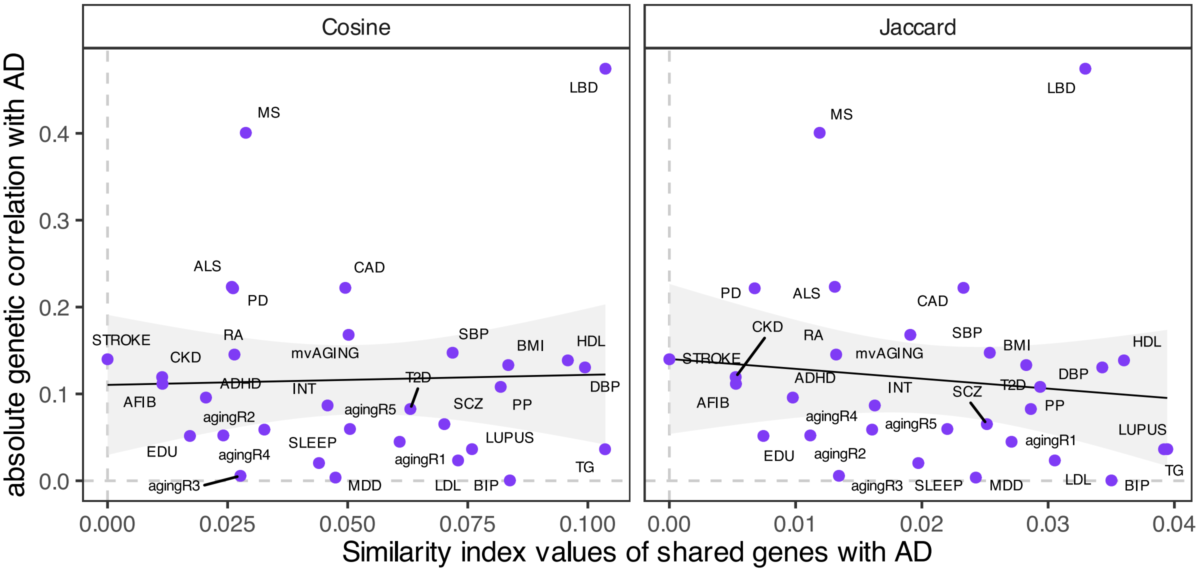

Displayed are scatterplots of cosine/Jaccard similarity indices (see **Methods** in the main text) and the absolute values of estimated genetic correlations between AD and each of the other 31 traits. Solid black lines are of the best fit through the purple points.

### Estimating SNP heritability of 32 traits

We estimated the SNP heritability of each of the 32 complex traits we analyzed in the main text using LD score regression (LDSC; Bulik-Sullivan et al., 2015) with the

European 1000 Genomes Phase 3 (Siva, 2008) reference panel and present the results in **Figure S4**. Standard errors were calculated using the principles of imputation (Rubin, 1996) across the 32 estimates made for each trait.

**Figure S4: LDSC-estimated SNP heritability for 32 traits**

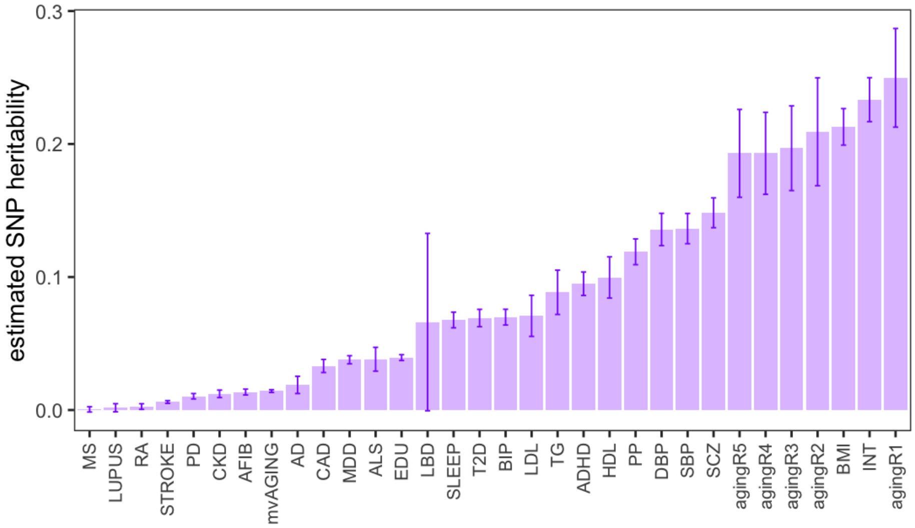

Displayed are estimates of SNP heritability and their 95% confidence interval using LD score regression (Bulik-Sullivan et al., 2015) and the European LD reference panel from 1000 Genome Phase 3 (Siva, 2008).

### Relationship between cosine/Jaccard index values and genetic correlation

**Figure S5** displays the association between squared genetic correlation and cosine/Jaccard index values for all pairs of the 32 traits we studied, subset to significant ($P<0.05$) genetic correlations. The linear correlation between significant squared genetic correlation and cosine index values is 0.16 ($P=0.018$), and with Jaccard index values is 0.17 ($P=0.011$).

**Figure S5: Cosine/Jaccard indices and genetic correlation for all 496 trait pairs**

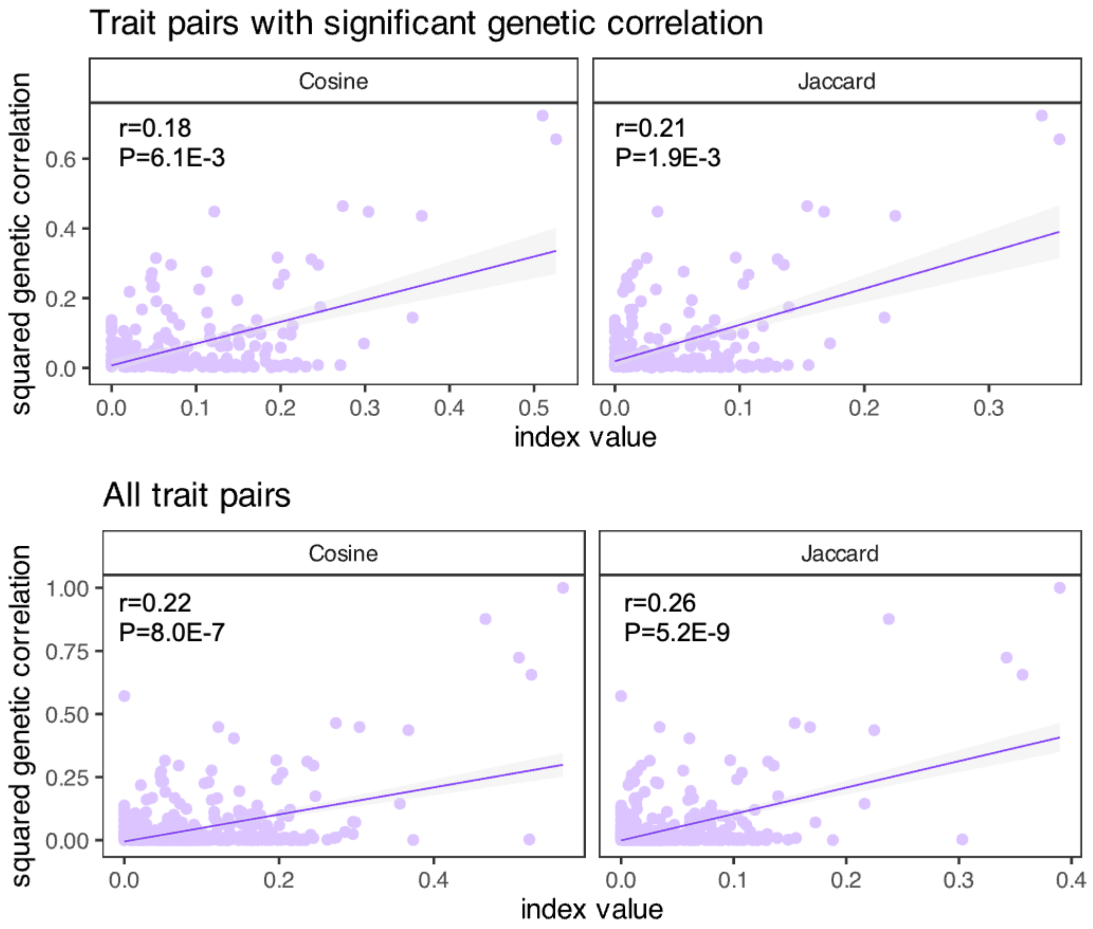

Displayed are scatterplots for cosine/Jaccard index values compared to squared estimated genetic correlations (LDSC; Bulik-Sullivan et al., 2015) for all 496 trait pairs. The top panel is restricted only to pairs with a genetic correlation that is considered 'significant' at the P<0.05 level. Purple lines are of best fit through the purple points. Pearson correlations are displayed in each panel above P-values that test the null hypothesis of no linear correlation.

### Adjusting shared causal gene counts for GWAS sample overlap

The algorithm we use to perform a simulation extrapolation adjustment to raw estimated counts of shared causal genes was introduced in the main text and the R code used to perform it in practice is available at https://github.com/noahlorinczcomi/bpact. We provide the original and adjusted estimates of shared causal gene counts between all pairs of traits in our full set of 32 in **Figure S6**. We truncated adjusted shared counts which were larger than original shared counts at the shared count value since the empirical evidence suggests adjusted shared counts should be less than or equal to the unadjusted/original shared counts. **Figure S6** shows that 94.8% of adjusted shared counts are equal to the original shared counts across all 496 pairs. Of the 26 pairs (5.2%) whose original and adjusted counts differed, the largest difference was of 5 genes, and the average difference was 1 gene. Together, these results suggest that the influence of GWAS sample overlap on estimated shared disease associated gene counts is not substantial in our analyses.

**Figure S6: Initial and adjusted estimated shared causal gene counts**

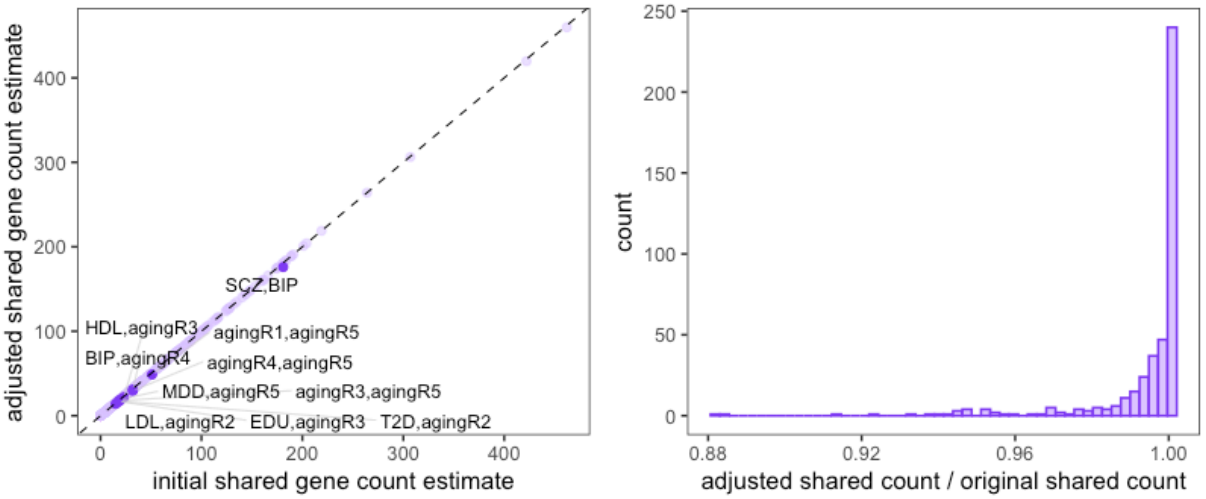

(left) Annotated points have adjustment factors less than 0.975 and original estimated shared gene counts greater than 10. (right) Distribution of the ratio of adjusted to original estimates of shared disease associated gene counts between all 496 trait pairs.

**Figure S7** displays results of the SIMEX adjustment for GWAS sample overlap applied to four pairs of traits: (i) DBP and SBP, (ii) MDD and BIP, (iii) SCZ and BIP, and (iv) SCZ and INT, each pair of which are phenotypically correlated and were studied in GWAS cohorts containing at least partially overlapping subjects. Subjects in the DBP and SBP GWAS were completely overlapping, so the displayed value of $\upsilon_{tt'}$, which was 0.78, is a direct estimate of their phenotypic correlation. The results in **Figure 7** show that for most pairs of traits increasing magnitudes of phenotypic correlation and GWAS sample overlap proportions (captured by the single term $\phi_{tt'}$) are associated with larger numbers of estimated shared disease associated genes, and that extrapolation from the simulated counts to the state of $\phi_{tt'} = 0$ reduces the original estimated counts of shared disease associated genes.

**Figure S7: Examples of SIMEX adjustment for GWAS sample overlap on shared causal gene counts**

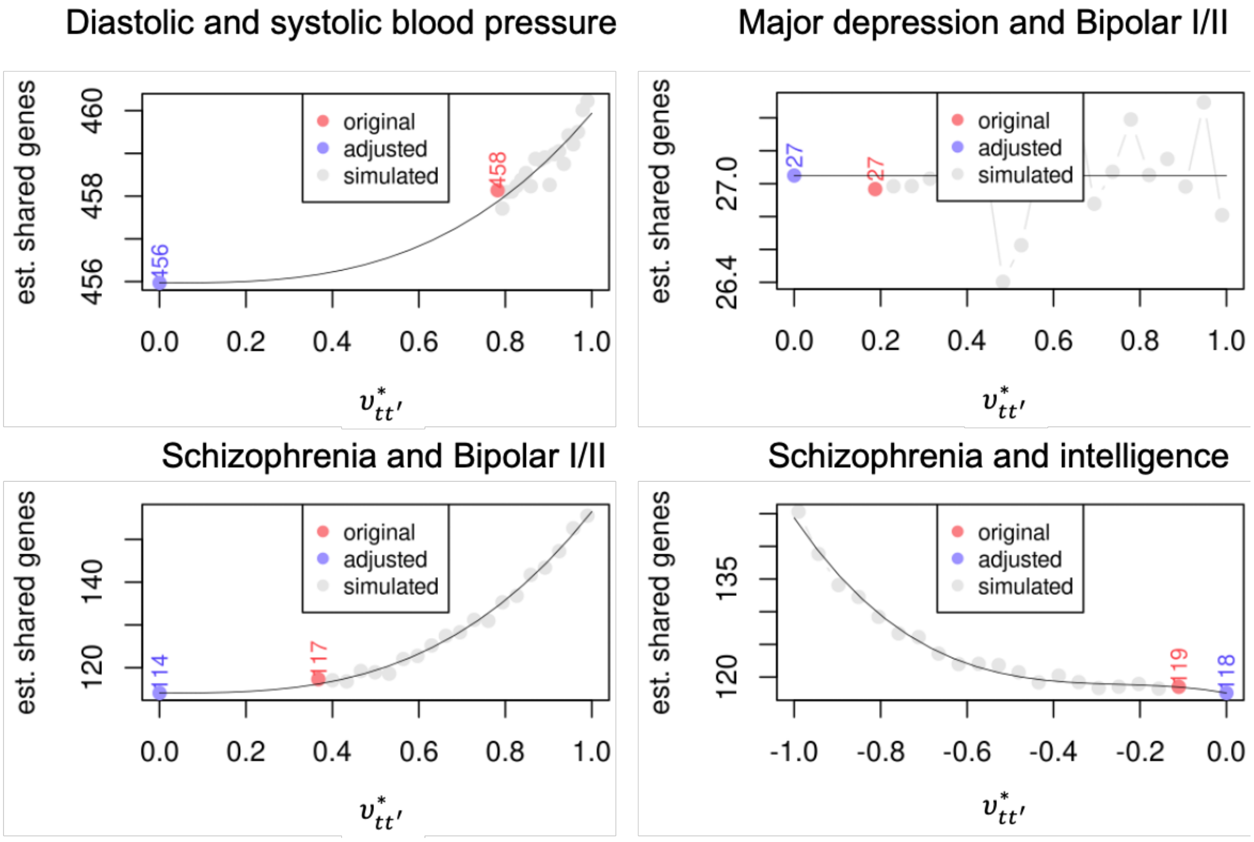

X-axis values represent the estimated phenotypic correlations between traits scaled by the proportion of overlapping GWAS subjects that was introduced in the main text. Y-axis values denote the estimated shared number of genes originally made by ignoring GWAS sample overlap (red point) and after adjusting for GWAS sample overlap (blue point) using extrapolation from simulation shared counts (grey points) under different scenarios of GWAS sample overlap using the SIMEX-type method introduced in the **Methods** section of the main text. Extrapolation was made using a third-order polynomial function fitted to all grey and red points that was penalized using elastic net regression with a regularization parameter that minimized cross-validation error and a ridge/LASSO tradeoff parameter of 0.5 (`glmnet` R package; Friedman et al., 2010).

### Additional supplementary figures and tables

**Figure S8: Correlation between estimated disease associated gene counts and GWAS sample size**

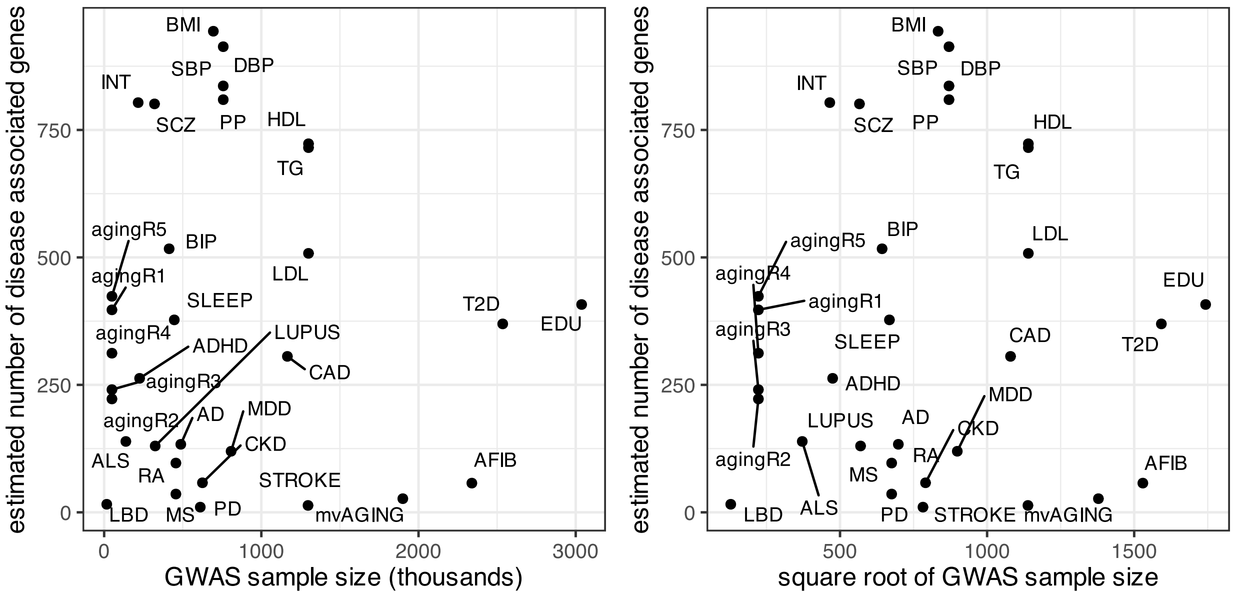

This figure is a scatterplot of the estimated number of disease-associated genes and the GWAS sample size for each trait. Estimated numbers of disease associated genes are made using summary level gene-based test statistics, which were calculated from SNP-level GWAS performed separately for each trait, and the estimator $c(t)$ from posterior causal probabilities introduced in the main text.

**Figure S9: Standardized sum of gene- and trait-specific posterior causal probabilities on each chromosome**

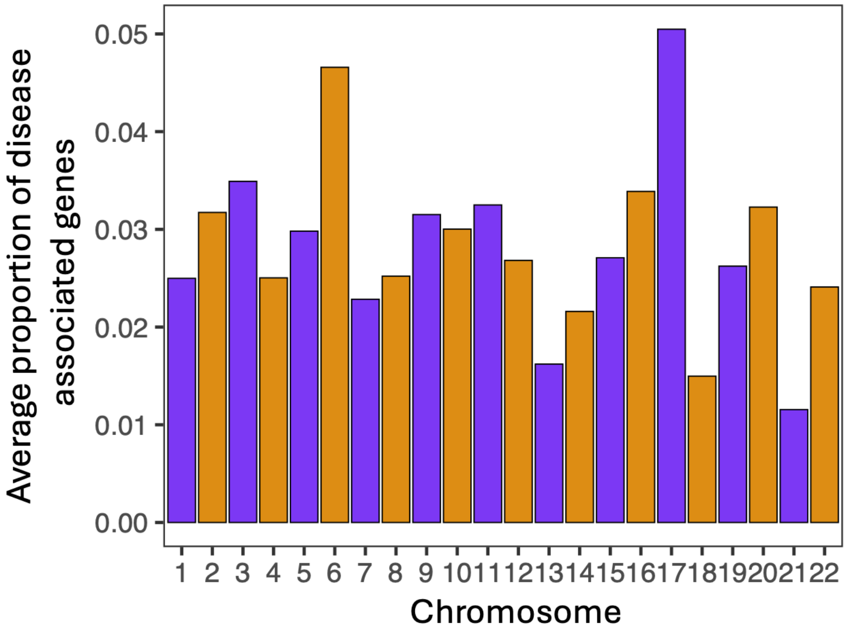

This figure displays, for each chromosome, the estimated number of disease-associated genes averaged across all traits divided by the total number of tested genes per trait. These results therefore reflect relative differences in estimated numbers of disease associated gene counts between chromosomes that are not influenced by larger numbers of tested genes on some chromosomes compared to others.

### Figure S10: Example pleiotropic and non-pleiotropic druggable gene targets for Alzheimer's disease

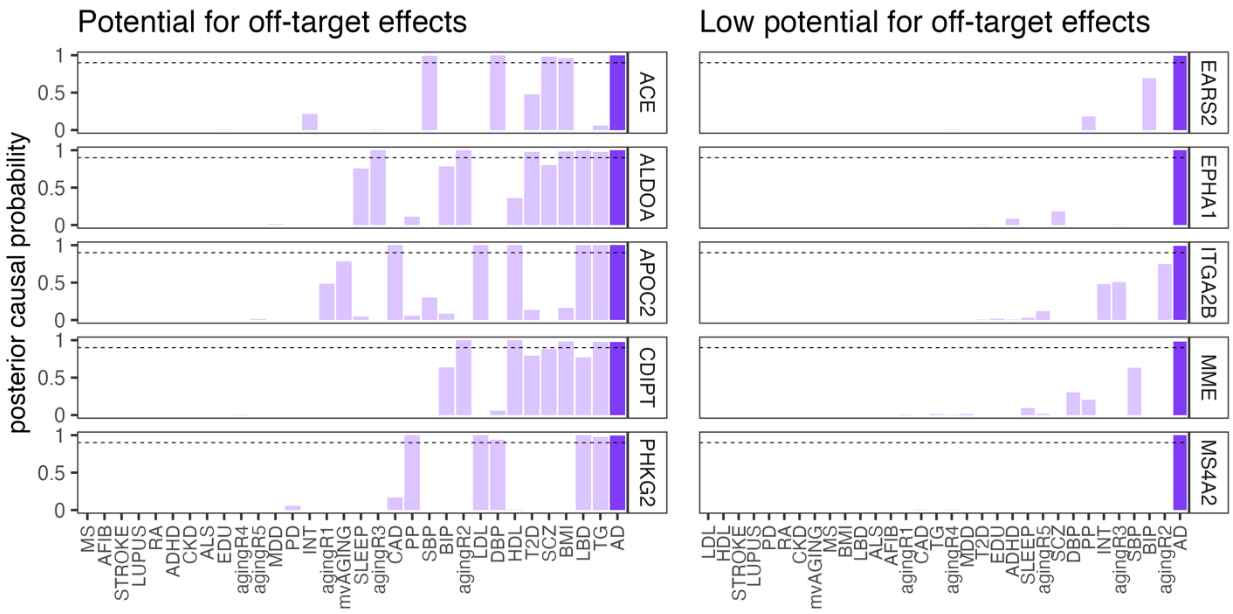

These results display posterior causal probabilities for 10 select genes with evidence of either a shared or non-shared association with any of the other 31 traits we studied.

**Figure S11: Genetic correlation estimates between all 32 traits (LDSC)**

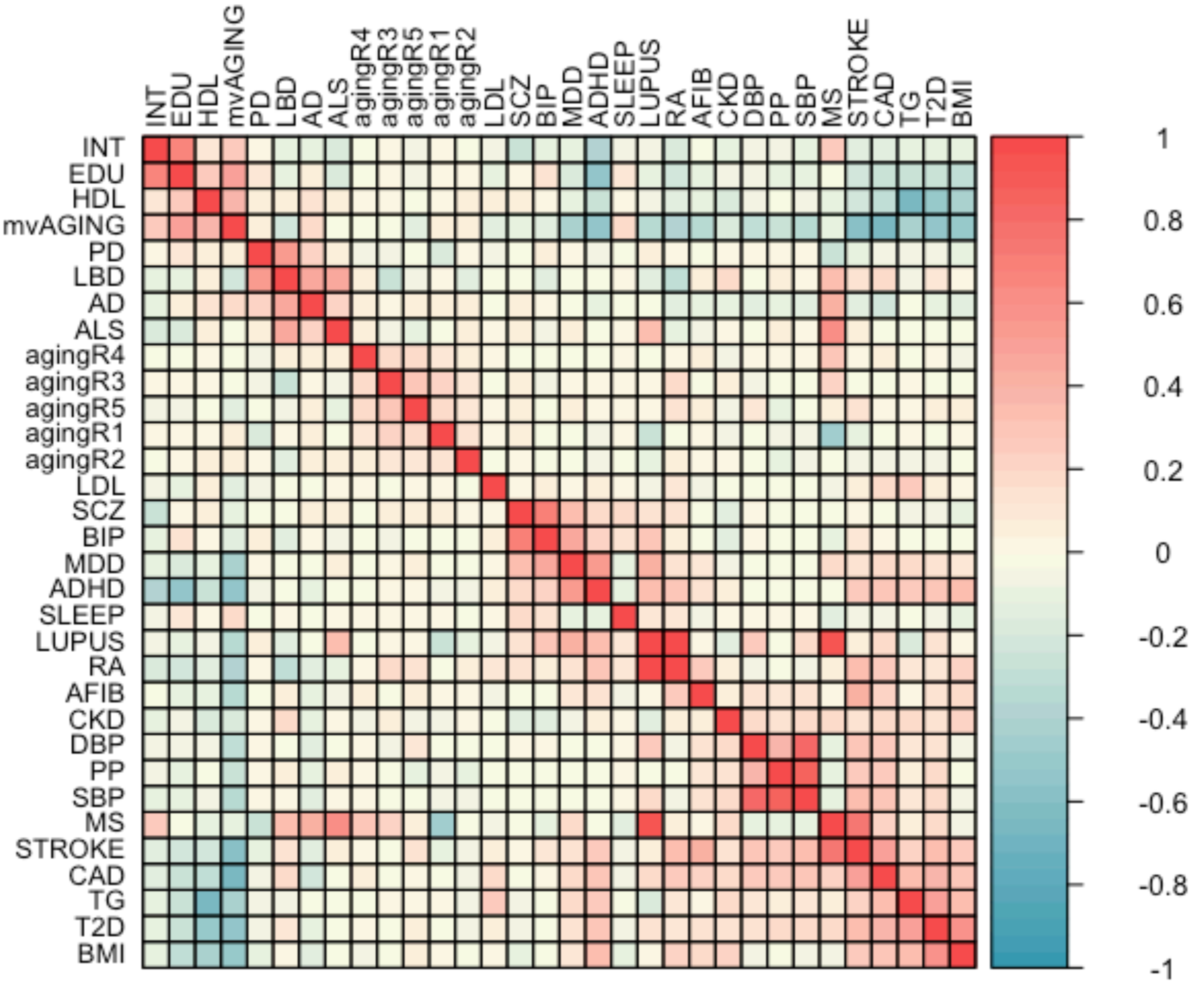

Displayed are estimates of genetic correlation between each pair of traits using LD score regression (Bulik-Sullivan et al., 2015). The order of traits on the rows and columns is determined using hierarchical clustering implemented in the `corrplot` R package (Wei & Simko, 2024).

**Supplementary Table S1: Run times (minutes) for steps in BPACT analysis**

| Composite likelihood Iteration | Posterior (trait 1) | Posterior (trait 2) | Proportion sharing (trait 1 and 2) | Adjusted proportion sharing (trait 1 and 2) |
|---|---|---|---|---|
| 1 | 4.99 | 4.93 | 1.75 | 1.05 |
| 2 | 5.11 | 5.35 | 2.10 | 1.03 |
| 3 | 5.58 | 5.93 | 2.84 | 1.03 |
| 4 | 5.02 | 4.89 | 1.70 | 1.05 |
| 5 | 4.97 | 4.96 | 1.84 | 1.04 |

Displayed are run times in minutes for the steps required to estimate the shared polygenic architecture between AD and LBD from 16,947 genes on an Intel® Xeon® Gold 6148 CPU @ 2.40GHz. ‘Posterior (trait1/2)’ refers to estimation of the posterior risk probability for each gene. ‘Proportion sharing (trait 1 and 2)’ refers to estimation of joint association between each gene and traits 1 (AD) and 2 (LBD). ‘Adjusted proportion sharing (trait 1 and 2)’ refers to the procedure which makes the SIMEX adjustment to the raw estimates of gene association sharing present in the ‘Proportion sharing (trait 1 and 2)’ column.

**References**

Borchers, H. W., & Borchers, M. H. W. (2019). Package ‘pracma’. *Practical numerical math functions, version*, *2*(5).

Bulik-Sullivan, B. K., Loh, P. R., Finucane, H. K., Ripke, S., Yang, J., Schizophrenia Working Group of the Psychiatric Genomics Consortium, ... & Neale, B. M. (2015). LD Score regression distinguishes confounding from polygenicity in genome-wide association studies. *Nature genetics*, *47*(3), 291-295.

Chib, S., & Greenberg, E. (1995). Understanding the metropolis-hastings algorithm. *The american statistician*, *49*(4), 327-335.

Friedman, J. H., Hastie, T., & Tibshirani, R. (2010). Regularization paths for generalized linear models via coordinate descent. Journal of statistical software, 33, 1-22.

Lorincz-Comi, N., Song, W., Chen, X., Rivera Paz, I., Hou, Y., Zhou, Y., Xu, J., Martin, W., Barnard, J., Pieper, A.A., Haines, J.L., Chung, M., & Cheng, F. (2024a). Combining xQTL and genome-wide association studies from ethnically diverse populations improves druggable gene discovery, *Cell Genomics*, under review.

Lorincz-Comi, N., Yang, Y., Li, G., & Zhu, X. (2024b). MRBEE: A bias-corrected multivariable Mendelian randomization method. Human Genetics and Genomics Advances, 5(3).

Rubin, D. B. (1996). Multiple imputation after 18+ years. Journal of the American statistical Association, 91(434), 473-489.

Siva, N. (2008). 1000 Genomes project. Nature biotechnology, 26(3), 256-257.

Wei T, Simko V (2024). R package 'corrplot': Visualization of a Correlation Matrix. (Version 0.95), https://github.com/taiyun/corrplot
